# Integrated immunovirological profiling validates plasma SARS-CoV-2 RNA as an early predictor of COVID-19 mortality

**DOI:** 10.1101/2021.03.18.21253907

**Authors:** Elsa Brunet-Ratnasingham, Sai Priya Anand, Pierre Gantner, Gaël Moquin-Beaudry, Alina Dyachenko, Nathalie Brassard, Guillaume Beaudoin-Bussières, Amélie Pagliuzza, Romain Gasser, Mehdi Benlarbi, Floriane Point, Jérémie Prévost, Annemarie Laumaea, Julia Niessl, Manon Nayrac, Gérémy Sannier, Marianne Boutin, Jade Descôteux-Dinelle, Gabrielle Gendron-Lepage, Guillaume Goyette, Catherine Bourassa, Halima Medjahed, Catherine Orban, Guillaume Butler-Laporte, David Morrison, Sirui Zhou, Tomoko Nakanishi, Laetitia Laurent, Jonathan Richard, Mathieu Dubé, Rémi Fromentin, Rose-Marie Rébillard, Nathalie Arbour, Alexandre Prat, Catherine Larochelle, Madeleine Durand, J Brent Richards, Michaël Chassé, Martine Tétreault, Nicolas Chomont, Andrés Finzi, Daniel E. Kaufmann

## Abstract

Despite advances in COVID-19 management, it is unclear how to recognize patients who evolve towards death. This would allow for better risk stratification and targeting for early interventions. However, the explosive increase in correlates of COVID-19 severity complicates biomarker prioritisation. To identify early biological predictors of mortality, we performed an immunovirological assessment (SARS-CoV-2 viral RNA, cytokines and tissue injury markers, antibody responses) on plasma samples collected from 144 hospitalised COVID-19 patients 11 days after symptom onset and used to test models predicting mortality within 60 days of symptom onset. In the discovery cohort (n=61, 13 fatalities), high SARS-CoV-2 vRNA, low RBD-specific IgG levels, low SARS-CoV-2-specific antibody-dependent cellular cytotoxicity, and elevated levels of several cytokines and lung injury markers were strongly associated with increased mortality in the entire cohort and the subgroup on mechanical ventilation. Model selection revealed that a three-variable model of vRNA, age and sex was very robust at identifying patients who will succumb to COVID-19 (AUC=0.86, adjusted HR for log-transformed vRNA=3.5; 95% CI: 2.0-6.0). This model remained robust in an independent validation cohort (n=83, AUC=0.85). Quantification of plasma SARS-CoV-2 RNA can help understand the heterogeneity of disease trajectories and identify patients who may benefit from new therapies.

## INTRODUCTION

Since the beginning of the pandemic, intense efforts have been deployed to define correlates of disease severity and to develop therapies targeting the virus or the pathogenesis of COVID-19. However, to date, only dexamethasone (*1–3*) and IL-6 blockers (tocilizumab (*4*), sarilumab (*5*)) have convincingly shown to provide a survival benefit in randomized controlled trials. While other immune interventions may benefit some subgroups (*6*), there is currently no consensus on how to predict which critical cases are likely to resolve their infection and which are at a greater risk of fatality, in part due to the high heterogeneity of patients and the very dynamic changes in biological features (*2*).

Recent reports have identified features linked to severe COVID-19. One is high amounts of viral RNA (vRNA) in plasma, which has been associated with greater severity and worst outcome for other respiratory pathogens, such as SARS-CoV-1 (*7, 8*), RSV (*9, 10*), MERS (*11*), and pandemic-causing strains of influenza A (H5N1 (*12*), H1N1 (*13*)). Plasma SARS-CoV-2 vRNA has also been linked with increased risk of severe COVID-19 and mortality (*14–17*).

Dysregulated immune responses are at least in part responsible for the exacerbated pathogenesis occurring in a minority of individuals with SARS-CoV-2 infection. Elevated cytokine levels were among the first reported markers associated to severe COVID-19 disease (*18*), although inconsistent sampling times sometimes led to weak associations with mortality (*19*). Narrowing the window of sampling early after symptom clarifies plasma cytokine patterns (*20*), reminiscent of the Cytokine Release Syndrome (*21*). Plasma profile around 10 days after symptom onset was highly differential for plasma cytokine profiles of critical *versus* moderate COVID-19 disease (*21*) and a number of cytokines have already been associated with increased mortality (*22*),

Multiple studies support a central role for antibody responses in protective anti-SARS-CoV-2 immunity. The main viral target of antibody immunity is the trimeric Spike glycoprotein, which facilitates SAR-CoV-2 entry into host cells via interaction of its receptor-binding domain (RBD) with angiotensin-converting enzyme-2 (ACE-2) (*23, 24*). While most infected patients develop anti-Spike and anti-RBD antibodies (*25, 26*), delayed anti-S IgG antibodies and decreased Fc-effector capacity are associated with increased mortality(*27*). These reports highlight the complexity of the host’s immune response to SARS-CoV-2.

Despite the remarkable speed with which effective SARS-CoV-2 vaccines have been developed and deployed, partial population coverage and, potentially, emergence of resistant variants will lead to ongoing occurrence of infections. From a clinical perspective, it is therefore essential to identify a minimal set of early blood parameters that can be easily and rapidly measured to identify patients at high risk of mortality, while prioritizing parameters that may hint at specific categories of therapeutic interventions. However, the list of blood correlates of COVID-19 severity has tremendously expanded, making such prioritizing a major challenge. Given strong co-upregulation between a number of plasma analytes (for example plasma cytokines and chemokines (*21*)), there is a need for simplified analytical models with few virological and/or immunological parameters that provide complementary, rather than redundant, information to better stratify individual patient risk.

In this study, we simultaneously examined multiple parameters in plasma spanning three key aspects of COVID-19 pathogenesis early in disease course (11 ± 4 days post symptom onset, henceforth described as DSO11): SARS-CoV-2 vRNA, 26 cytokines and tissue injury markers, and 6 measures of SARS-CoV-2-specific antibody responses. We performed uni- and multivariable analysis to identify independent predictors of death. A minimal model combining vRNA, age and sex was particularly robust, and very reproducible in an independent validation cohort.

## RESULTS

### Study design and patient characteristics

We investigated prospectively enrolled hospitalized COVID-19 individuals (n=144) with symptomatic infection and a positive SARS-CoV-2 nasopharyngeal swab PCR. These patients were infected during the first wave, and were not enrolled in immunotherapy trials. Our study population was split into a discovery cohort (n=61) in a first hospital and a fully independent validation cohort (n=83) in a second hospital (see Study Design, Figure S1A, and participant characteristics, Table 1). To allow for cross-sectional analysis of early plasma markers, we investigated patients for whom research blood samples were available at 11 (± 4) days after symptom onset (DSO11). Based on disease severity at DSO11, patients were grouped as critical (requiring mechanical ventilation) *versus* non-critical. The discovery cohort included 29 critical and 32 non-critical patients. Plasma profiles were compared to 50 asymptomatic uninfected donors as a control group (uninfected controls – UC) of non-diseased state.

**Table 1:**
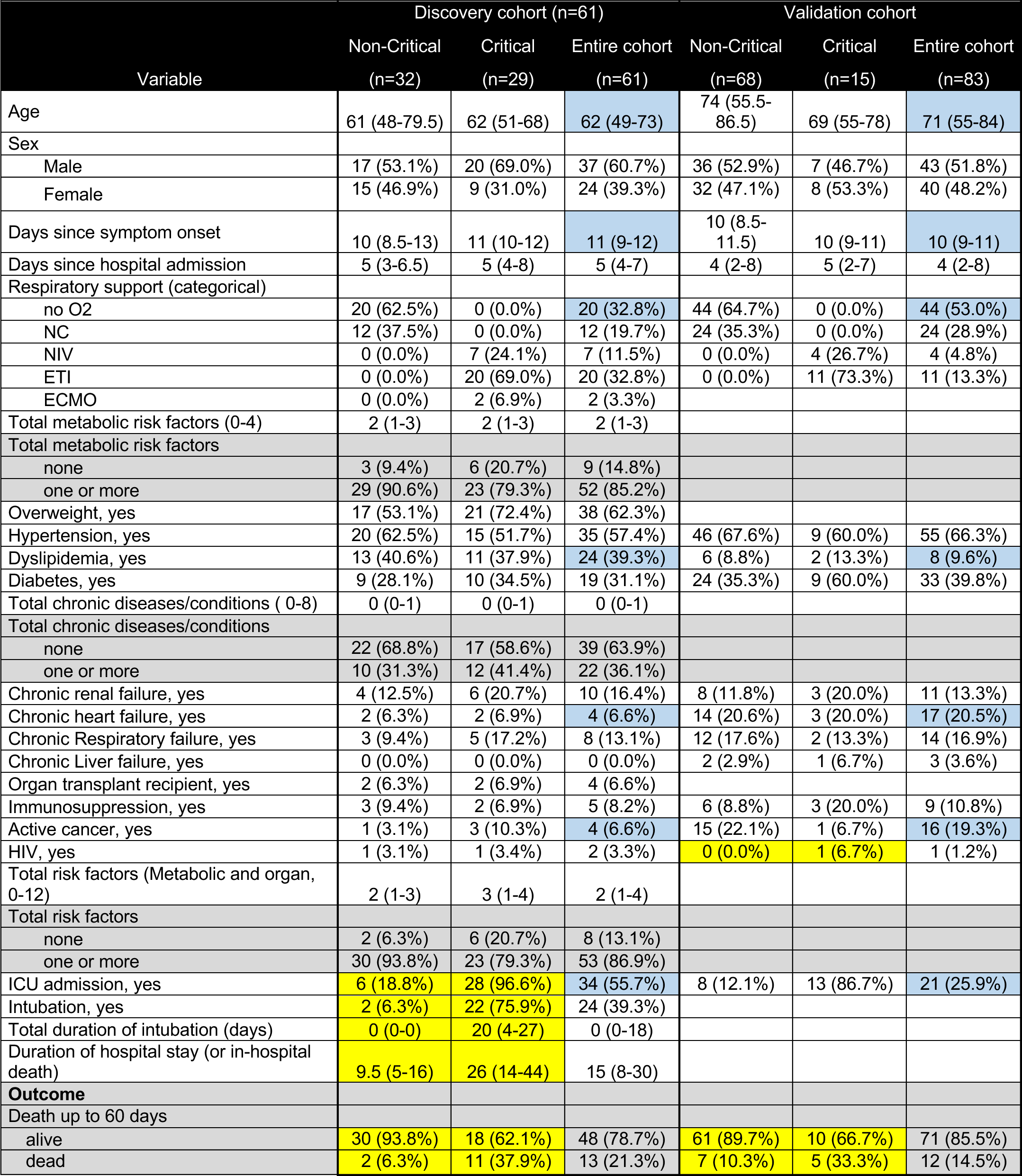
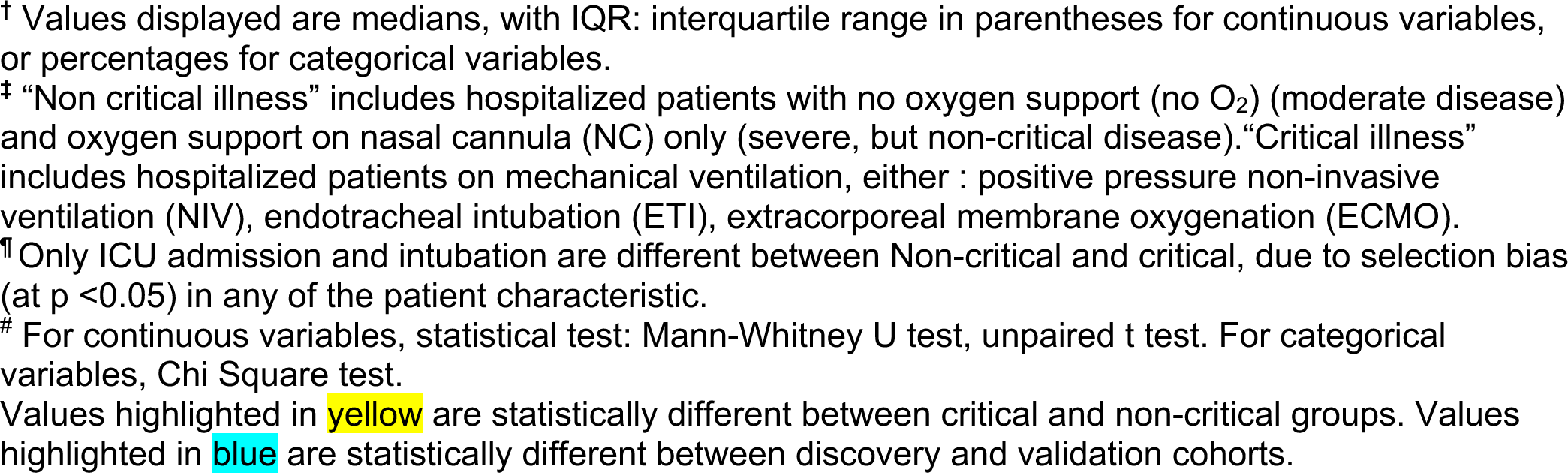
Baseline characteristics of the participants and respiratory support at time of immunovirological profiling. ^†^.

| Variable | Discovery cohort (n=61) |  |  | Validation cohort |  |  |
| --- | --- | --- | --- | --- | --- | --- |
|  | Non-Critical<br>(n=32) | Critical<br>(n=29) | Entire cohort<br>(n=61) | Non-Critical<br>(n=68) | Critical<br>(n=15) | Entire cohort<br>(n=83) |
| Age | 61 (48-79.5) | 62 (51-68) | 62 (49-73) | 74 (55.5-86.5) | 69 (55-78) | 71 (55-84) |
| Sex |  |  |  |  |  |  |
| Male | 17 (53.1%) | 20 (69.0%) | 37 (60.7%) | 36 (52.9%) | 7 (46.7%) | 43 (51.8%) |
| Female | 15 (46.9%) | 9 (31.0%) | 24 (39.3%) | 32 (47.1%) | 8 (53.3%) | 40 (48.2%) |
| Days since symptom onset | 10 (8.5-13) | 11 (10-12) | 11 (9-12) | 10 (8.5-11.5) | 10 (9-11) | 10 (9-11) |
| Days since hospital admission | 5 (3-6.5) | 5 (4-8) | 5 (4-7) | 4 (2-8) | 5 (2-7) | 4 (2-8) |
| Respiratory support (categorical) |  |  |  |  |  |  |
| no O2 | 20 (62.5%) | 0 (0.0%) | 20 (32.8%) | 44 (64.7%) | 0 (0.0%) | 44 (53.0%) |
| NC | 12 (37.5%) | 0 (0.0%) | 12 (19.7%) | 24 (35.3%) | 0 (0.0%) | 24 (28.9%) |
| NIV | 0 (0.0%) | 7 (24.1%) | 7 (11.5%) | 0 (0.0%) | 4 (26.7%) | 4 (4.8%) |
| ETI | 0 (0.0%) | 20 (69.0%) | 20 (32.8%) | 0 (0.0%) | 11 (73.3%) | 11 (13.3%) |
| ECMO | 0 (0.0%) | 2 (6.9%) | 2 (3.3%) |  |  |  |
| Total metabolic risk factors (0-4) | 2 (1-3) | 2 (1-3) | 2 (1-3) |  |  |  |
| Total metabolic risk factors |  |  |  |  |  |  |
| none | 3 (9.4%) | 6 (20.7%) | 9 (14.8%) |  |  |  |
| one or more | 29 (90.6%) | 23 (79.3%) | 52 (85.2%) |  |  |  |
| Overweight, yes | 17 (53.1%) | 21 (72.4%) | 38 (62.3%) |  |  |  |
| Hypertension, yes | 20 (62.5%) | 15 (51.7%) | 35 (57.4%) | 46 (67.6%) | 9 (60.0%) | 55 (66.3%) |
| Dyslipidemia, yes | 13 (40.6%) | 11 (37.9%) | 24 (39.3%) | 6 (8.8%) | 2 (13.3%) | 8 (9.6%) |
| Diabetes, yes | 9 (28.1%) | 10 (34.5%) | 19 (31.1%) | 24 (35.3%) | 9 (60.0%) | 33 (39.8%) |
| Total chronic diseases/conditions ( 0-8) | 0 (0-1) | 0 (0-1) | 0 (0-1) |  |  |  |
| Total chronic diseases/conditions |  |  |  |  |  |  |
| none | 22 (68.8%) | 17 (58.6%) | 39 (63.9%) |  |  |  |
| one or more | 10 (31.3%) | 12 (41.4%) | 22 (36.1%) |  |  |  |
| Chronic renal failure, yes | 4 (12.5%) | 6 (20.7%) | 10 (16.4%) | 8 (11.8%) | 3 (20.0%) | 11 (13.3%) |
| Chronic heart failure, yes | 2 (6.3%) | 2 (6.9%) | 4 (6.6%) | 14 (20.6%) | 3 (20.0%) | 17 (20.5%) |
| Chronic Respiratory failure, yes | 3 (9.4%) | 5 (17.2%) | 8 (13.1%) | 12 (17.6%) | 2 (13.3%) | 14 (16.9%) |
| Chronic Liver failure, yes | 0 (0.0%) | 0 (0.0%) | 0 (0.0%) | 2 (2.9%) | 1 (6.7%) | 3 (3.6%) |
| Organ transplant recipient, yes | 2 (6.3%) | 2 (6.9%) | 4 (6.6%) |  |  |  |
| Immunosuppression, yes | 3 (9.4%) | 2 (6.9%) | 5 (8.2%) | 6 (8.8%) | 3 (20.0%) | 9 (10.8%) |
| Active cancer, yes | 1 (3.1%) | 3 (10.3%) | 4 (6.6%) | 15 (22.1%) | 1 (6.7%) | 16 (19.3%) |
| HIV, yes | 1 (3.1%) | 1 (3.4%) | 2 (3.3%) | 0 (0.0%) | 1 (6.7%) | 1 (1.2%) |
| Total risk factors (Metabolic and organ, 0-12) | 2 (1-3) | 3 (1-4) | 2 (1-4) |  |  |  |
| Total risk factors |  |  |  |  |  |  |
| none | 2 (6.3%) | 6 (20.7%) | 8 (13.1%) |  |  |  |
| one or more | 30 (93.8%) | 23 (79.3%) | 53 (86.9%) |  |  |  |
| ICU admission, yes | 6 (18.8%) | 28 (96.6%) | 34 (55.7%) | 8 (12.1%) | 13 (86.7%) | 21 (25.9%) |
| Intubation, yes | 2 (6.3%) | 22 (75.9%) | 24 (39.3%) |  |  |  |
| Total duration of intubation (days) | 0 (0-0) | 20 (4-27) | 0 (0-18) |  |  |  |
| Duration of hospital stay (or in-hospital death) | 9.5 (5-16) | 26 (14-44) | 15 (8-30) |  |  |  |
| **Outcome** |  |  |  |  |  |  |
| Death up to 60 days |  |  |  |  |  |  |
| alive | 30 (93.8%) | 18 (62.1%) | 48 (78.7%) | 61 (89.7%) | 10 (66.7%) | 71 (85.5%) |
| dead | 2 (6.3%) | 11 (37.9%) | 13 (21.3%) | 7 (10.3%) | 5 (33.3%) | 12 (14.5%) |

<sup>†</sup> Values displayed are medians, with IQR: interquartile range in parentheses for continuous variables, or percentages for categorical variables.
<sup>‡</sup> “Non critical illness” includes hospitalized patients with no oxygen support (no O<sub>2</sub>) (moderate disease) and oxygen support on nasal cannula (NC) only (severe, but non-critical disease). “Critical illness” includes hospitalized patients on mechanical ventilation, either : positive pressure non-invasive ventilation (NIV), endotracheal intubation (ETI), extracorporeal membrane oxygenation (ECMO).
<sup>¶</sup> Only ICU admission and intubation are different between Non-critical and critical, due to selection bias (at $p < 0.05$ ) in any of the patient characteristic.
<sup>#</sup> For continuous variables, statistical test: Mann-Whitney U test, unpaired t test. For categorical variables, Chi Square test.
Values highlighted in yellow are statistically different between critical and non-critical groups. Values highlighted in blue are statistically different between discovery and validation cohorts.

We clinically followed participants for at least 60 days after symptom onset (DSO60). The primary outcome, death by DSO60, occurred in 13 patients (21.3%), with close to half fatalities occurring between DSO30 and DSO60 (Figure S1B) and mostly in the critical group (Figure S1C).

We performed a more focused immunovirological assessment in the validation cohort, where 15 cases were critical, and 68 non-critical, and 12 deaths occurred before DSO60. Because of hospital referral coordination, the validation cohort was older than the discovery one, but with less severe respiratory compromise (Table 1). Other basic demographics and prevalent risk factors were consistent with published studies(*28*) and overall showed minor differences between both cohorts. These features were also not different between the critical vs non-critical groups except for higher rates of admission to ICU and intubation, and duration of hospital stay in critical patients (Table 1), in line with group definition.

### Plasma viral load in early disease is strongly associated with COVID-19 mortality

As SARS-CoV-2 vRNA in plasma has been previously linked to mortality, we quantified it in the discovery cohort. We designed an ultrasensitive quantitative real time PCR (qRT-PCR) targeting the N sequence of its genome with a detection limit of 13 copies/mL. The assay was highly specific, with no vRNA detected in UC (Figure 1A). At DSO11, we detected plasma SARS-CoV-2 vRNA in a significantly greater fraction of critical than non-critical patients (76% vs 28%, Figure 1A). These results suggest that systemic SARS-CoV-2 viremia is a signature of infection severity and/or itself plays a role in disease complications.

**Figure 1.**
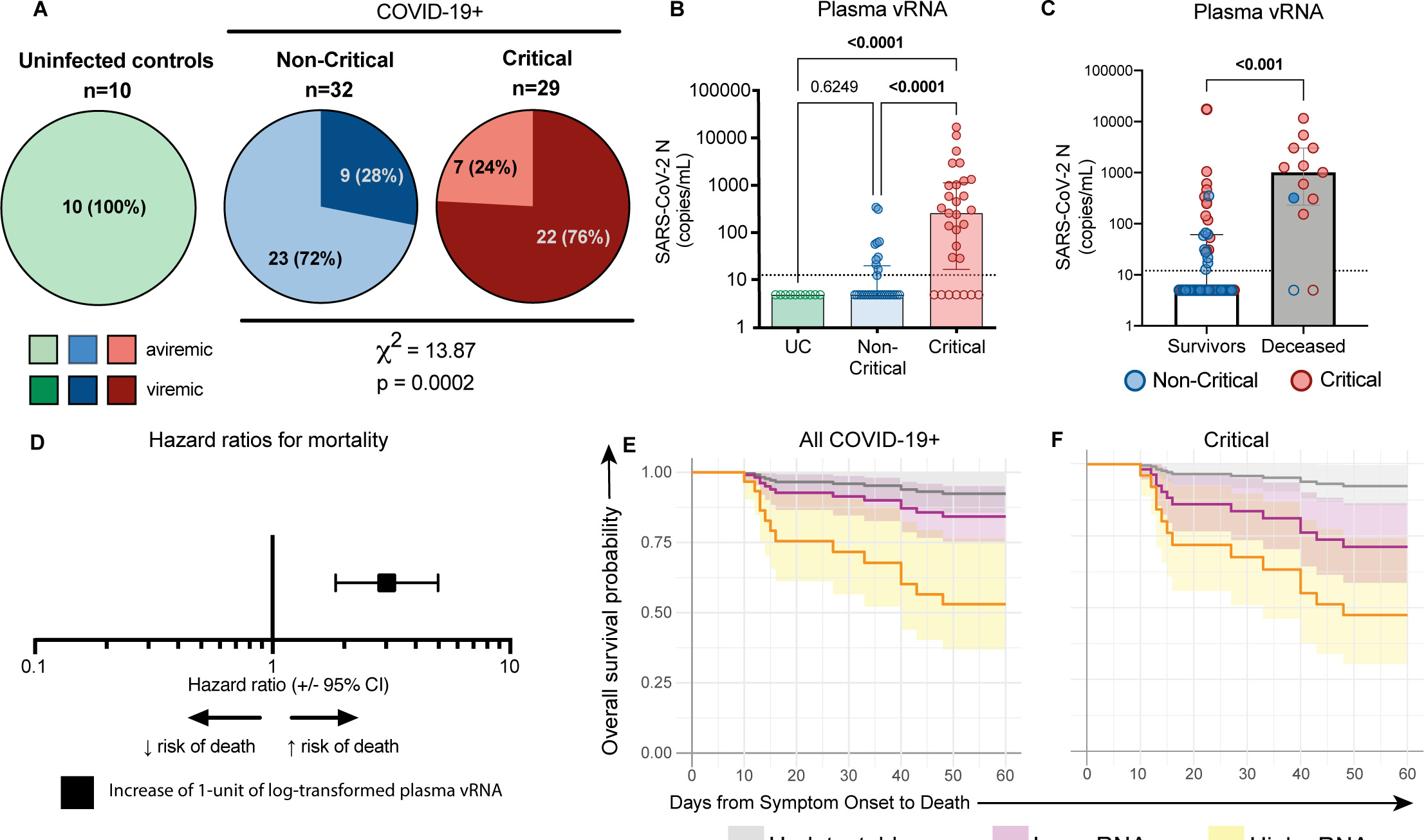
Plasma SARS-CoV-2 RNA levels at DSO11 are associated with increased risk of mortality. **A)** Pie charts representing the fractions of assessed samples which had undetectable (aviremic, light shades, <13 copies/mL) or detectable SARS-CoV-2 vRNA (*≥*13 copies/mL, dark shades). Numbers in parts refer to the number (and percentages) of patients within each cohort/group. Non-critical and critical groups compared by Chi Square test. **B)** Quantities of N copies of SARS-CoV-2 RNA detected per mL of plasma in each cohort/group. Dotted line is the limit of detection (13 copies/mL). Empty shapes have undetectable vRNA (arbitrarily set at 5 copies/mL for representation). **C)** Amounts of N copies of SARS-CoV-2 RNA detected per mL of plasma in patients who survived (white column) or died (grey column) by DSO60. Red circles represent critical patients, and blue are non-critical. **D)** HR with 95%CI calculated using Cox regression for an increase of 1 unit of log_10_-transformed vRNA (copies/mL). **EF**) Modelization of the hazard ratio of patients with high (orange, upper IQR), low (purple, lower IQR) or undetectable (grey) plasma vRNA in **E)** all COVID-19 patients or **F)** Critical cases only. B) Kruskall-Wallis with Dunn’s multiple comparisons test. C) Mann-Whitney test. n = 61 COVID-19 subjects (13 mortalities) or 29 Critical COVID-19 cases (11 mortalities) and 10 UC. IQR = interquartile range, calculated among detectable vRNA quantities only.

We next hypothesized that the amount of viral products, rather than their mere presence, was associated with severe pathogenesis. SARS-CoV2 vRNA levels were higher in critical than non-critical cases (Figure 1B). This difference held when the comparison was restricted to samples with detectable plasma vRNA (p=0.002 - Mann-Whitney test). Most patients who died had high vRNA compared to survivors (Figure 1C). In univariate Cox regression analysis (Table 1. 2) we found that an increase of 1 unit of log-transformed plasma vRNA led to a 3-fold increase in mortality risk [Hazard Ratio (HR)= 3.0 (95% Confidence Interval CI: 1.8-5.0), p < 0.0001 for all COVID-19 (Figure 1D), and 2.4 (95% CI: 1.3 – 4.4, p=0.006) for the critical group (Table 2)]. The estimated survival proportions for undetectable (<13 copies/ml), low, or high plasma vRNA were extracted from Cox models (see methods for details) (*29*). High plasma vRNA was associated with a greater risk of death, whereas there was substantial overlap between the subgroups with low or undetectable plasma vRNA (Figure 1E). A similar trend was observed in the critical group (Figure 1F). Therefore, plasma SARS-CoV-2 vRNA load is not only a correlate of contemporaneous respiratory compromise early in disease course, but is also associated with mortality, including in the critical group.

**Table 2.** Univariate Cox proportional hazard regression of single variables measured in COVID-19 patient plasma at DSO11.

| Variable | Discovery cohort |  |  |  |
| --- | --- | --- | --- | --- |
|  | All COVID-19+ at DSO11 (n=61) |  | Critical COVID-19+ subset at DSO11 (n=29) |  |
|  | HR(95%CI)<br>1 unit | p value | HR(95%CI)<br>1 unit | p value |
| **Viral Load** |  |  |  |  |
| vRNA (copies/mL of plasma)* | 3.0 (1.8, 5.0) | <0.001 | 2.4 (1.3, 4.4) | 0.006 |
| **Serology** |  |  |  |  |
| RBD-specific IgG (RLU)* | 0.3 (0.1, 0.8) | 0.011 | 0.3 (0.1, 0.7) | 0.005 |
| RBD-specific IgM (RLU)* | 0.5 (0.2, 1.4) | 0.186 | 0.4 (0.1, 1.3) | 0.144 |
| RBD-specific IgA (RLU)* | 0.4 (0.2, 0.98) | 0.045 | 0.3 (0.1, 0.8) | 0.014 |
| Spike Ig (MFI)* | 0.6 (0.5, 0.9) | 0.006 | 0.5 (0.3, 0.8) | 0.002 |
| Neutralization (ID50)* | 0.8 (0.6, 1.1) | 0.172 | 0.7 (0.5, 0.9) | 0.020 |
| ADCC (%) ♦ | 0.7 (0.5, 0.9) | 0.006 | 0.1 (0.5, 0.9) | 0.004 |
| **Cytokines** |  |  |  |  |
| Angiopoietin-2* | 14.5 (3.4, 62.1) | 0.001 | 6.7 (1.4, 33.0) | 0.018 |
| CCL2/JE/MCP-1* | 5.6 (1.7, 18.4) | 0.005 | 2.8 (0.8, 10.2) | 0.115 |
| CCL20/MIP-3 alpha* | 2.9 (1.2, 6.8) | 0.016 | 1.4 (0.5, 4.0) | 0.578 |
| CCL7/MCP-3/MARC* | 4.0 (1.0, 15.6) | 0.050 | 5.2 (0.9, 30.7) | 0.068 |
| CD40 Ligand/TNFSF5* | 5.6 (1.0, 30.8) | 0.049 | 6.7 (0.8, 55.7) | 0.080 |
| CXCL10/IP-10/CRG-2* | 16.7 (0.7, 423.5) | 0.088 | 5.5 (0.2, 161.2) | 0.323 |
| CXCL13/BLC/BCA-1* | 6.7 (2.6, 17.2) | <0.001 | 4.3 (1.6, 11.7) | 0.005 |
| IL-8/CXCL8* | 5.6 (1.7, 18.6) | 0.005 | 3.8 (1.0, 14.3) | 0.048 |
| CXCL9/MIG* | 2.2 (0.8, 6.4) | 0.133 | 1.2 (0.5, 2.9) | 0.621 |
| D-dimer* | 5.0 (0.5, 49.9) | 0.174 | 0.4 (0.02, 8.5) | 0.548 |
| G-CSF* | 3.3 (1.1, 10.2) | 0.034 | 3.5 (1.1, 10.8) | 0.032 |
| GM-CSF* | 9.4 (1.7, 50.7) | 0.009 | 7.3 (1.02, 51.4) | 0.047 |
| IFNα* | 2.4 (0.9, 6.5) | 0.087 | 2.4 (0.8, 7.0) | 0.114 |
| IL-1ra/IL-1F3* | 8.0 (1.9, 33.7) | 0.004 | 2.8 (0.6, 14.4) | 0.214 |
| IL-23* | 13.7 (2.2, 85.6) | 0.005 | 7.2 (1.1, 46.3) | 0.038 |
| IL-6* | 2.6 (1.4, 5.0) | 0.003 | 1.5 (0.7, 3.3) | 0.315 |
| SP-D* | 5.2 (1.0, 26.3) | 0.047 | 2.2 (0.3, 15.7) | 0.433 |
| TNFα* | 16.5 (2.7, 102.5) | 0.003 | 6.6 (0.9, 50.9) | 0.069 |
| RAGE/AGER* | 7.9 (2.3, 27.9) | 0.001 | 4.4 (1.01, 18.9) | 0.049 |
| CytoScore** | 2.6 (1.6, 4.2) | <0.001 | 1.9 (1.1, 3.4) | 0.0335 |

\*Variables are log<sub>10</sub> transformed. HR shown are for an increase of 1 unit of log<sub>10</sub>-transformed variable.
\*\*Refer to Material and Methods for details.
♦HR for increase of 10 units.
RLU = Relative Light Units, normalized to internal control (CR3022) (see methods for details). ADCC = antibody-dependent cellular cytotoxicity.

### Markers of immune hyperactivation and tissue damage discriminate disease trajectories

As early elevation of a number of cytokines and chemokines was also associated with adverse COVID-19 outcome (*20, 21, 30*), we used multiplexed beads arrays to determine plasma levels of 26 proteins associated with adaptive and/or innate immune responses, chemotaxis, or tissue insult related to severe acute respiratory distress syndrome (ARDS, See Table S1 for analyte list). Principal component analysis revealed that the plasma profile largely delineates UC from COVID-19 patients, and highlighted higher cytokine levels and greater heterogeneity in the critical group compared to the non-critical group (Figure 2A) The outlier critical case at the upper left corner of the PCA was on extracorporeal membrane oxygenation (ECMO) at the time of sampling, a procedure known to affect plasma profile(*31*). Unsupervised hierarchical clustering parsed apart 3 patient clusters: I) mostly critical; II) mixed; III) mostly non-critical cases (Figure 2B).

**Figure 2.**
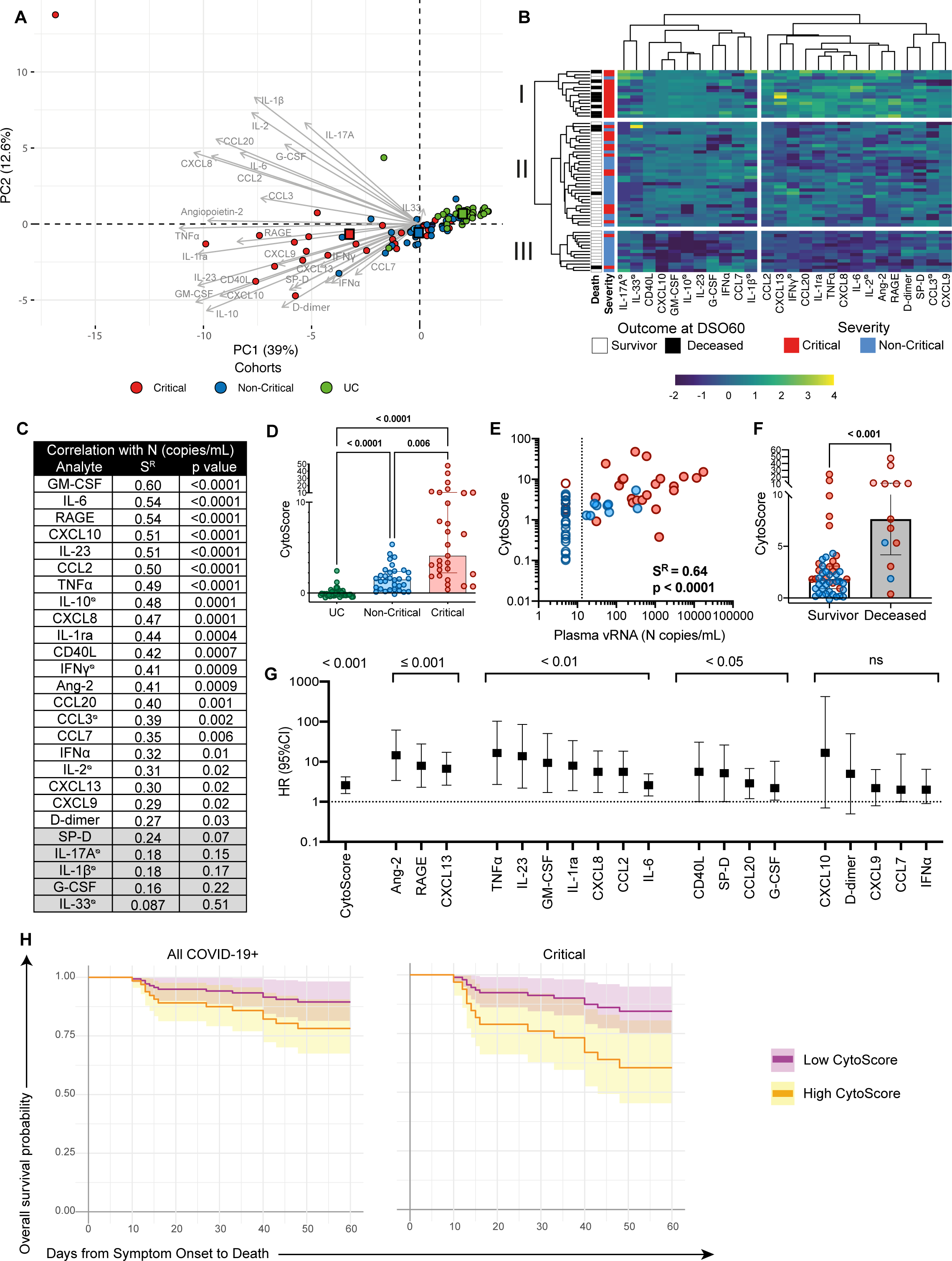
High cytokine titers in plasma at DSO11 discriminate critical disease and is associated with increased risk of mortality. **A)** Principal component analysis (PCA) representation of critical and non-critical patients (at DSO11), and UC (at baseline), on the basis of the 26 plasma analytes. Color-coded squares represent the mean PC coordinates for each group. Length of arrow indicates contribution of analytes to PCs. Numbers in parentheses along axes are the percentage of variance that PC accounts for. **B)** Heatmap analysis of log-transformed concentrations of all 26 plasma analytes (yellow = high relative expression; blue = low relative expression), with unsupervised hierarchical clustering of the analytes (top dendrogram) or of patients (left dendrogram). Left-most column represents outcome at DS60 (white = survival, black = deceased). Following column is the severity of the patient at DSO11. **C**) Table showing the Spearman R values and corresponding p values of correlation of each plasma analyte with plasma vRNA. Values shaded in grey are non-significant. **D)** Comparison of CytoScore of each cohort (see methods for details on CytoScore). **E)** Correlation between plasma vRNA and CytoScore. Empty shapes are aviremics (<13 copies SARS-CoV-2/mL of plasma). **F)** CytoScore of patients whom survived (white column) or deceased (grey column) by DSO60. **G**) HR with 95%CI calculated using Cox regression for a 1-unit increase of the log_10_-transformed concentration of each plasma analyte with robust detection (see methods for details) and CytoScore. **HI)** Modelization of the hazard ratio of patients with high (orange, upper IQR) or low (purple, lower IQR) CytoScore in **H)** all COVID-19 patients or **I)** Critical subgroup only. CE) Spearman correlations. D) Kruskall-Wallis with Dunn’s multiple comparisons test. F) Mann-Whitney test. For A-B, D-F, color-coded dots represent severity of the patient at DSO11 (red = critical, blue = non-critical), or UC cohort (green). BD) Cytokines with titles annotated by ⊘ are poorly detected (See methods for details). n = 61 COVID-19 subjects (13 mortalities) or 29 Critical COVID-19 cases (11 mortalities) and 43 UC. IQR = interquartile range, calculated within the CytoScores of the COVID-19 discovery cohort.

We next compared the levels of each analyte between groups (Figure S2A-D). Several followed a stepwise increase, where non-critical cases had greater cytokine concentrations than UC, and critical cases had the greatest amounts (Figure S2A). These included pro-inflammatory cytokines such as IL-6, GM-CSF and TNF*α* and pro-inflammatory chemokines CCL2 and CXCL8. Some of the markers of tissue insult (RAGE, Angiopoietin-2)(*32*) also increased with disease severity, likely reflecting the extent of lung and vascular damage. CXCL9, CD40L, IFN*α* and surfactant pulmonary protein D (SP-D) were significantly greater only in the critical cases of COVID-19 compared to UC (Figure S2B), while a few markers did not differ between all three groups (Figure S2C). Some analytes were significantly elevated in COVID-19 groups but did not differ between the critical and non-critical groups, such as the chemokines CXCL10 (IP10) and CXCL13, and D-dimer (Figure S2D). Taken together, the plasma profile reveals overall higher quantities of cytokines in the plasma of COVID-19 patients compared to UC, and select analytes are specifically associated with greater disease severity.

We reasoned these 26 analytes may be differentially linked to the amount of vRNA in plasma. We examined the correlations between individual plasma analytes (Figure S2E), as well as their association with vRNA (Figure 2C). Many analytes were co-upregulated, and several of them also positively correlated with vRNA levels. These latter correlations were particularly robust for cytokines implicated in innate immune responses such as IL-6 (Figure S2F) and GM-CSF, the marker of endothelial damage RAGE (Figure S2G), and inflammatory chemokines CXCL8, CXCL10, and CCL2, suggesting a shared trigger or overlap in pathways.

To capture by a single parameter the overall magnitude of the difference in cytokine titers between COVID-19 patients and UC, we created a “CytoScore” from the linear combination of all 26 analytes (See methods for details). CytoScore followed a gradual difference, where the non-critical group had lower CytoScores than critical, and UC had the lowest scores (Figure 2D). The CytoScore correlated positively with vRNA (Figure 2E) and can have value as a way to reduce dimensionality of plasma analyte profiling.

As patients who died within DSO60 showed a greater CytoScore than survivors (Figure 2F), we applied Cox regressing analyses to examine the association between the cytokines and mortality over time. We focused on analytes whose concentrations are in the range of robust quantitation by the assay (19/26, see methods for details). For each, we calculated the HR associated with a 1-unit increase of log-transformed concentration (Figure 2G). Several individual analytes were significantly associated with increased fatality risk, with Angiopoientin-2, RAGE and CXCL13 showing the highest significance (p ≤ 0.001). As the CytoScore was also highly significant, we compared the predicted survival probability of patients with low or high CytoScore at DSO11 (Figure 2H). The latter population showed a significantly lower rate of predicted survival at DSO60 than the low CytoScore population. This observation was maintained when we restricted our analysis to the critical group (Figure 2I). Therefore, overall cytokine levels as well as individual cytokines and markers of tissue damage measured at DSO11 are 1) in majority correlated with plasma vRNA and 2) associated with increased risk of mortality among COVID-19 patients.

### Low SARS-CoV-2-specific IgG and limited ADCC responses are associated with COVID-19 mortality

As SARS-CoV-2 antibody responses likely play a critical role in protective immunity against SARS-CoV-2 (*27, 33*), we measured plasma SARS-CoV-2-specific antibody responses at DSO11. ELISA-based quantification using the SARS-CoV-2 RBD protein and isotype-specific secondary antibodies (*25, 34*) revealed a broad range in relative quantities of RBD-specific IgM, IgA or IgG in the non-critical and critical groups at DSO11. They did not differ between groups, and were not detected in UC (Figure 3A). These observations were corroborated by a flow cytometry-based assay measuring plasma binding to full-length Spike protein (Spike Ig) on cell surface (Figure 3B), which similarly showed no significant difference between the two COVID-19 groups.

**Figure 3.**
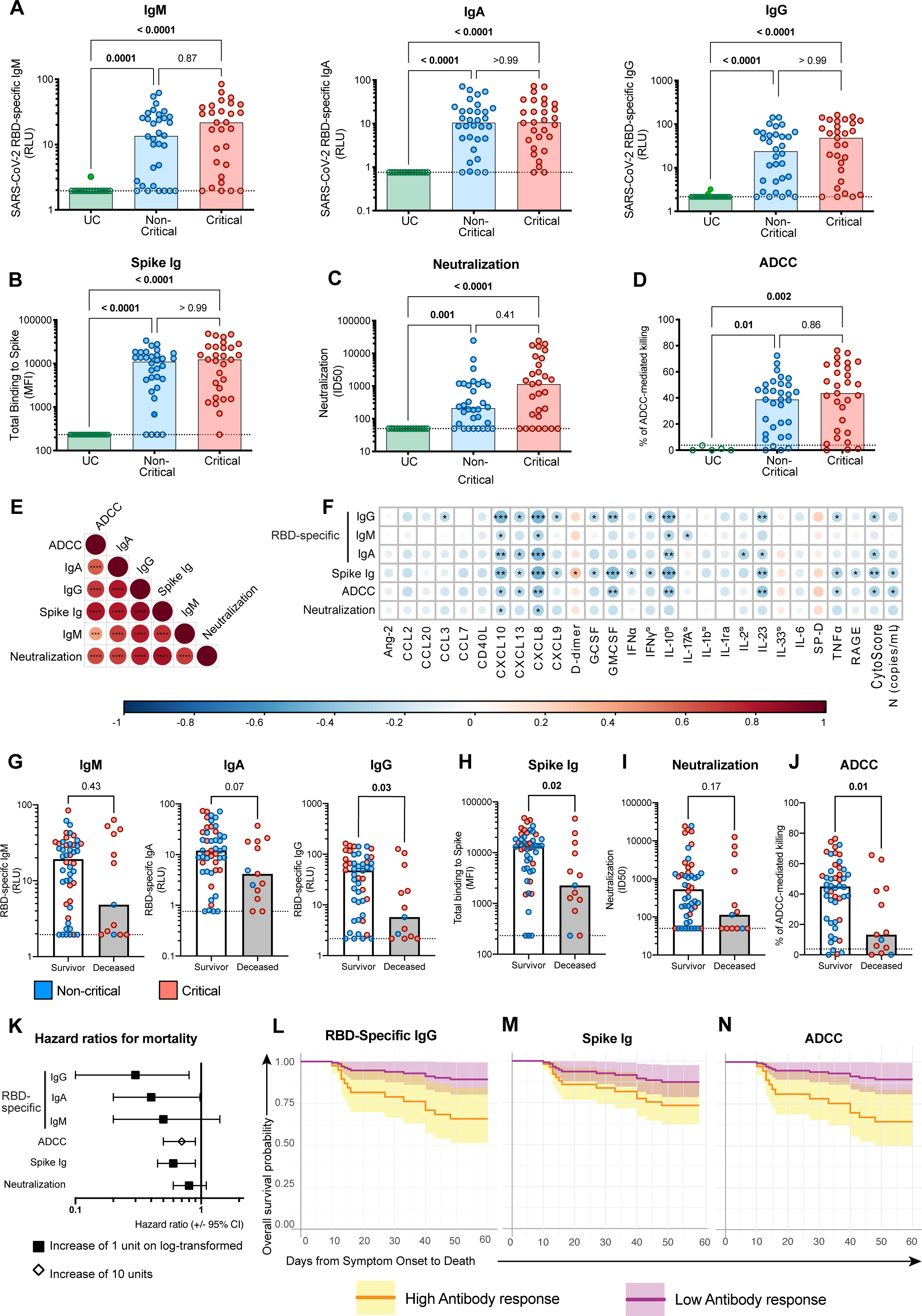
Limited SARS-CoV-2-specific IgG responses and low ADCC at DSO11 are associated with mortality. **A)** ELISA-based relative quantification of SARS-CoV-2 RBD-specific antibodies’ isotypes IgM (left), IgA (center) or IgG (right) in Relative light units (RLU) normalized to an internal control (CR3022). **B-D)** Comparison of functional properties of the plasma of all three groups, namely **B**) plasma capacity to recognize the SARS-CoV-2 full Spike (Spike Ig) using a flow cytometry-based assay (Median Fluorescence Intensity - MFI); **C**) plasma neutralization activity (unit = half of maximal inhibitory plasma dilution – ID50); **D**) plasma ADCC activity (unit = % of ADCC-mediated killing). **EF)** Correlation matrixes with colors representing the Spearman R value (blue = negative association -1; red = positive association 1), and p values indicated as * in the circles, **E)** between all serology measurements or **F)** of serology measurements *versus* plasma vRNA and plasma analytes. **G-J)** Comparison of serology measurements in patients who survived (white column) or deceased (grey column) by DSO60, for **G)** RBD-specific IgM (left), IgA (center) or IgG (right) or **H)** Full Spike binding, **I**) Neutralization or **J**) ADCC. **K**) Hazard ratio with 95%CI calculated using Cox regression for an increase of 1 unit of log_10_-transformed (square) or 10 units (diamond) of serology measurements. **L-N**) Modelization of the hazard ratio over time of patients with high (orange, upper IQR) or low (purple, lower IQR) **L**) RBD-specific IgG, **M**) Spike Ig or **N**) ADCC activity in all COVID-19 patients. A-D) Kruskall-Wallis with Dunn’s multiple comparison test. EF) Spearman R correlation. F) Cytokines with titles annotated by ⊘ are poorly detected. G-J) Mann-Whitney test. For G-J, color-coded dots represent severity of the patient at DSO11 (red = critical, blue = non-critical) and dotted line represents the limit of detection. EFL) p <0.05 = *; p <0.01 = ***; p < 0.001 = ***. n = 61 COVID-19 subjects (13 mortalities) or 29 Critical COVID-19 cases (11 mortalities) and 43 UC. IQR = interquartile range, calculated within the COVID-19 discovery cohort.

We next assessed the SARS-CoV-2 Spike-specific antibody response for two key antiviral functions: neutralization (Figure 3C) and antibody-dependent cellular cytotoxicity (ADCC, Figure 3D). Here again, the data showed high variability, and no significant differences between the critical and non-critical groups for both readouts. All serology measurements were interrelated (Figure 3E). In contrast, the serology measurements were inversely correlated with plasma vRNA and most cytokines (Figure 3F).

To assess potential consequences of defective antibody responses at this early time point, we compared SARS-CoV-2-specific antibody responses between survivors and non-survivors. For RBD-specific isotypes (Figure 3G), only IgG amounts were significantly increased in survivors, although there was a similar trend for IgA as well. Spike Ig levels were also higher in survivors (Figure 3H). We observed contrasting patterns with regard to functional humoral responses: while neutralization capacity was similar for the two outcomes (Figure 3I), ADCC capacity was superior in survivors (Figure 3J). HR reflected the same observations, where higher ADCC, RBD-specific IgG and Spike Ig were associated with increased survival (Figure 3K). We further modeled this by comparing the survival curves at DSO60 of patients with low or high RBD-specific IgG amounts (Figure 3L), Spike Ig (Figure 3M) or ADCC (Figure 3N) at DSO11, and saw that participants with low responses for these three measurements showed an increased fatality risk. These observations were maintained when the analysis was restricted to the critical group (Figure S3A-C). Taken together, these results highlight that impairment of some SARS-CoV-2-specific antibody responses may contribute to mortality.

### Multivariate Cox survival models of early immunovirological features reveal a pivotal role for plasma vRNA association with COVID-19 mortality

As all categories of immunovirological parameters showed some perturbations that predicted fatality, we examined whether these alterations provided redundant information in terms of mortality risk, or if their combined analysis would improve associations with fatal outcome. Within immunovirological categories, we retained only variables significant in univariate Cox analysis (p < 0.05; see Table 2), and among those, a global multivariate model was used to select top variables (See methods for details). To evaluate predictive accuracy of the resulting variables and multivariate models, time-dependent receiver operator characteristic (ROC) curves were calculated at DSO60 (principles illustrated in Figure 4A, see methods for details). The area under the curve (AUC), a measure of prediction accuracy, was examined at all distinct event times by plotting the AUC curve over time (principles illustrated in Figure S5A, see methods for details). All final Cox models were reassessed in the validation cohort, and their time dependent ROC curves were evaluated to validate the accuracy of our findings.

**Figure 4.**
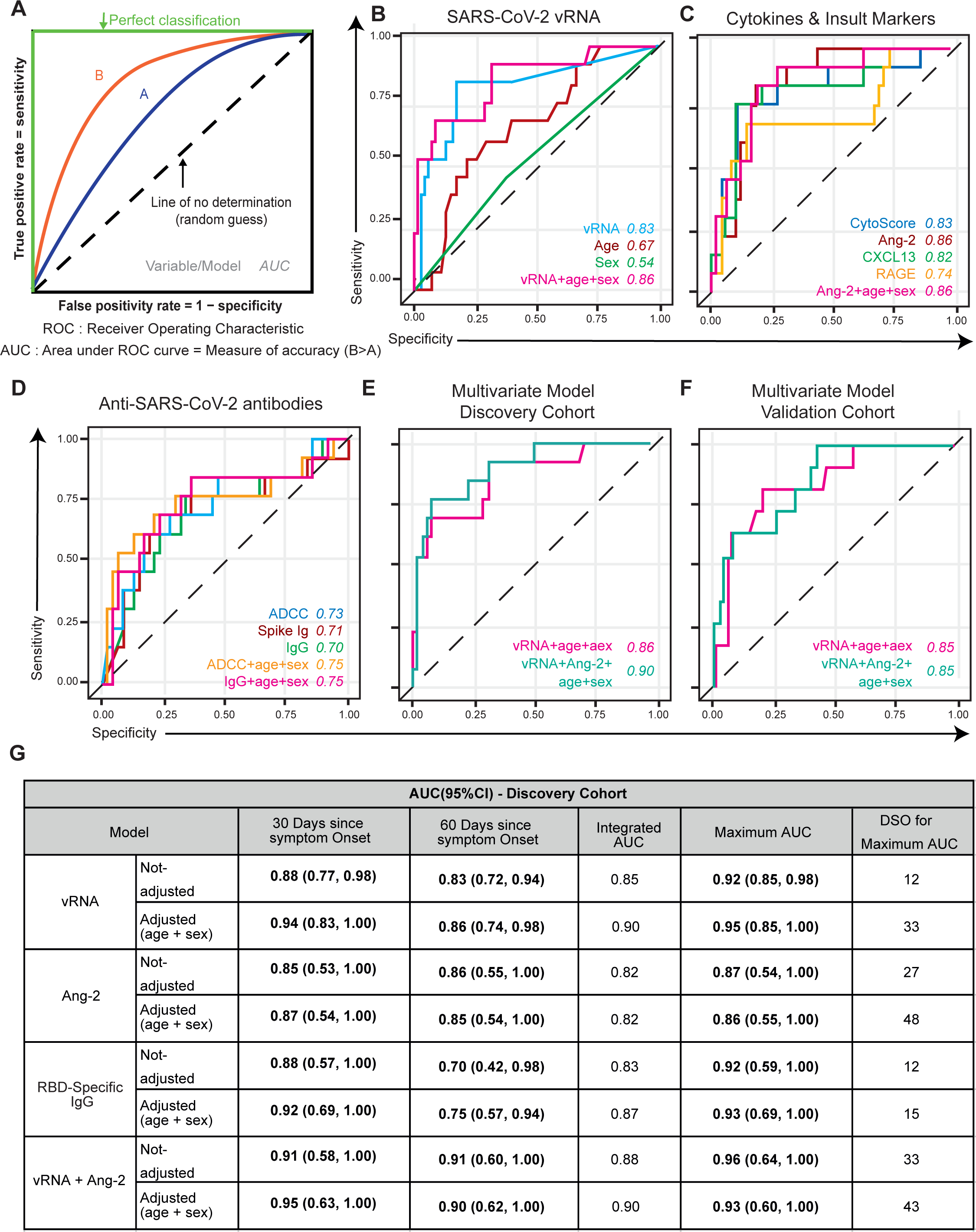
Time-dependent ROC curves reveal plasma vRNA as reproducibly associated with mortality in both the discovery and validation cohorts. **A**) Concept of time-dependent ROC curves. A ROC is defined by the false-positive rate and true-positive rate, which depicts relative trade-offs resulting from changing the test threshold. The best possible prediction model (100% sensitivity and 100% specificity) would yield a “square curve”, reaching the upper left corner (green line). A completely random guess (chance) would give a point along the diagonal dotted black line (line of no determination). In the present study, the ROC curves were used compare the predictive accuracy of different immunovirological parameters measured in plasma at DSO11. These ROC curves were time-dependant, meaning they vary depending on the time between symptom onset and death considered. Here, model B is superior to A. ROC curves are further characterized by AUC, a measure of test accuracy (1.0= best possible test; 0.5=no discrimination). **B-D)** Time-dependent ROC curves measured within the discovery cohort for **B)** plasma vRNA, age and sex; **C)** Cytokines and tissue insult markers or **D)** anti-SARS-CoV-2 antibody responses. **E-F)** Time-dependent ROC curves of top multivariate models selected by BIC stepwise selection in the **E**) Discovery cohort, and their validation in the **F**) Validation cohort. **G**) Table summarizing time-dependent AUC for representative variables per category in discovery cohort. Values are AUC (95%CI), and AUC at p<0.05 are in bold. AUC values given either not adjusted (only variable(s) listed) or adjusted (age and sex). AUC values are given at 30 days or 60 days after symptom onset. Maximum AUC column gives the best prediction accuracy of the variable (measured at DSO11), and DSO for maximum is the date since symptom onset at which that maximum AUC was achieved. B-F) Legends with color-coded variables are on the bottom left of panels, and values in italic are the AUC values associated to the variable. n = 61 for discovery cohort; 83 for validation cohort.

In the discovery cohort, time-dependent ROC for plasma vRNA showed a strong predictive capacity at DSO60 (AUC=0.83, 95%CI: 0.72-0.94), and a slight benefit when adjusting for age and sex (AUC=0.86, 95%CI:0.74-0.98) (Figure 4B). The AUC for the adjusted model reached a high of 0.95 (Figure 4G) at DSO33 but mostly hovered around 0.90 (Figure S5B). When applied to the validation cohort at DSO60, vRNA again had a good predictive capacity (AUC=0.76; 95%CI:0.58-0.95), and a benefit when adjusting for age and sex (AUC=0.85; 95%CI: 0.71-1.00) (Figure S4A). The highest AUC of the adjusted model (0.92; 95%CI:0.85-1.00) was reached at DSO22 (Figure 4G), but was stable over time (Figure S5C). Therefore, vRNA is a strong predictor of fatality, and adjusting for age and sex improves its predictive power.

Next, we compared the time-dependent ROC curves for inflammatory and tissue damage markers of the discovery cohort (Figure 4C). Multivariate model selection retained only 1 analyte: Angiopoientin-2. To compare predictive accuracies at DSO60, we selected 3 additional analytes highly significant (p ≤ 0.001) in univariate Cox (Figure 2G): CytoScore, CXCL13 and RAGE (Figure 4C). Although no individual inflammatory cytokine was selected, the CytoScore had a high AUC (0.83, 95%CI: 0.71-0.95). Of the two markers of tissue insult, only Angiopoietin-2 (AUC=0.86; 95%CI: 0.55-1.00) remained significant (RAGE: AUC=0.74; 95%CI: 0.33-1.00). The chemokine CXCL13 (AUC=0.82, 95%CI: 0.70-0.94) also had good predictive accuracy. All AUC values stayed quite stable over time (Figure S5D). When applied to the validation cohort, only CXCL13’s remained high (AUC=0.82, 95%CI: 0.66-0.98) and significantly discriminatory of mortality (p<0.05) (Figures S4DE) over time (Figure S5E). These observations confirm that certain markers of tissue insult and chemokine, as well as the overall cytokine levels, were associated with mortality risk.

For antibody measurements, we observed, within the discovery cohort, overlap of the time-dependent ROC of all three measurements significant in univariate Cox (ADCC: AUC=0.73, 95%CI: 0.44-1.00; Spike Ig: AUC=0.71, 95%CI: 0.25-1.00; RBD-specific IgG: AUC=0.70, 95%CI:0.42-0.98) at DSO60 (Figure 4D, G). However, the predictive value of all 3 measurements began to drop around DSO30 (Figure S5F). We then applied the analysis to the validation cohort. The cell-based ADCC assay requires significant infrastructure and technical expertise that may not be available in all clinical settings. We therefore removed ADCC from the validation list of variables, leading to its substitution by the technically simple RBD-specific IgG, in line with their strong correlation (Figure 3C). The time-dependent ROC curves in the validation cohort for Spike Ig (AUC=0.62; 95%CI: 0.15-1.00) and RBD-specific IgG titers (AUC=0.61, 95%CI: 0.17-1.00) were non-significant, and lower than in the discovery cohort. However, they displayed good predictive accuracy of mortality with DSO60 when adjusted for age and sex (Spike Ig: 0.78, 95%CI: 0.54-1.00; RBD-specific IgG: 0.78, 95%CI: 0.53-1.00) (Figure S4C). Taken together, these data reveal that the anti-SARS-CoV-2 antibody response is highly associated with mortality within 30 days of symptom onset, but less so afterwards.

After examining each variable in the setting of their category, we sought to identify which single parameter, or combination thereof, is the most robust. All variables selected by multivariate model within each category were considered for a global multivariate model, and age and sex covariates were forced regardless of their significance. In the discovery cohort, the variables selected in the global multivariate model (AUC=0.90, 95%CI: 0.62-1.00) were vRNA (HR = 2.47, 95%CI:1.30-4.68) and Angiopoietin-2 (HR = 4.22, 95%CI: 0.66-26.78), alongside the forced variables age (HR = 1.06, 95%CI: 0.99-1.10) and sex (HR= 0.94, 95%CI: 0.24,3.70) (Figure 4E). Only vRNA (p=0.006) remained independently associated with a higher risk of all-cause mortality within DSO60 in the global multivariate model. The predictive accuracy of the global multivariate model remained quite stable overtime (Figures 4G, S5H). In the validation cohort, the it’s predictive accuracy was non-significant (AUC:0.85, 95%CI: 0.32-1.00) (Figures 4FG). However, the exclusion of Angiopoietin-2 improved the model’s discrimination in the validation cohort; this three-variable model combining vRNA, age and sex was then significant (AUC=0.85; 95%CI: 0.71-1.00) and stable over time (Figure S5I).

Taken together, these data indicate that, at DSO11, measuring plasma SARS-CoV-2 vRNA in hospitalised COVID-19 patients can be a powerful tool to predict mortality.

## DISCUSSION

In the perspective of clinical translation, it is essential to rigorously select among the multitude of markers linked to COVID-19-related mortality. In patients with a spectrum of disease severity, we studied perturbations within three categories of plasma molecules associated with death: circulating SARS-CoV-2 vRNA (*16*), elevated immune and tissue injury markers (*30*) and inadequate SARS-CoV-2-specific antibody responses (*27*), all of which can be probed by quick and technically robust assays. Strong associations of early parameters with our primary outcome, fatality within 60 days of symptom onset, were observed, and largely maintained when the analyses were restricted to the critical group of patients on mechanical ventilation. Multivariate analyses demonstrated that, because of collinearity between several variables, a limited number of biological features was sufficient to build robust models predicting mortality. stand out as an early feature critically, and consistently, associated with higher mortality risk. Combined analysis of SARS-CoV-2 vRNA, Angiopoietin-2, age and sex had greatest predictive accuracy in a discovery cohort, although a simpler model with vRNA, age and sex was almost as robust. This three-parameter model maintained significant predictive accuracy in an independent validation cohort. In our study, plasma vRNA levels therefore stand out as an early feature critically, and consistently, associated with higher mortality risk.

The strength of the association between plasma vRNA levels and mortality risk was stronger than previously reported for nasopharyngeal swabs (NSW) (*35*). In contrast to plasma, quantification of vRNA in NSW is hard to normalize, varies between types of tests, and depends on sample quality. Cox models showed a 3-fold increase of fatal outcome for every 1-unit increase in log-transformed plasma vRNA quantity. While this association is reminiscent of the remarkable predictive value of plasma viral load for disease progression in untreated HIV-1 infection (*36*), no study has thus far convincingly demonstrated that therapeutic reduction of SARS-CoV-2 viral loads decreased mortality risk. For example, the antiviral remdesivir reduced viral loads in NSW, duration of symptoms, and hospitalization, but had no significant impact on survival (*37, 38*).

Similarly, although monoclonal anti-Spike antibodies can reduce viral load (*39, 40*), trials have not yet shown benefit in hospitalized patients. Given disease heterogeneity, it will therefore be important to determine if such interventions specifically benefit the subgroup of patients with high plasma vRNA.

The source and precise nature of the plasma vRNA remains to be better determined. Viral nucleic acids in the plasma do not prove the presence of replication-competent viral particles, as they could be viral debris translocated from damaged lung tissue. This is supported by the correlation we saw between vRNA and RAGE. Whereas the increase in plasma of RAGE’s ligand EN-RAGE was linked to its’ increased mRNA expression in the PBMCs of severe COVID-19 patients, RAGE mRNA was not expressed in this compartment (*30*), supporting that plasma RAGE originates directly from damaged tissue (*32*). Besides the direct cytopathic effects of SARS-CoV-2 on lung epithelium, immunopathological mechanisms are thought to play a key role in severe COVID-19 pathogenesis (*41*). Systemic vRNA may trigger pathogen-recognition receptors such as TLRs, in line with strong co-upregulation of interferon-stimulated genes (ISGs) and other inflammatory pathways in vRNA-containing cells (*42*). This could contribute to the strong correlation observed between the amount of vRNA and IL-6, a pathogen-associated molecular pattern (PAMP)-triggered inflammatory cytokine (*43*).

Consistent with previous studies (*20, 21*), we found significant associations between levels of several immune and tissue damage markers with both disease severity and mortality. Despite strongly significant HR for fatality risk for some analytes, the small sample size of our study resulted in sizeable overlaps between confidence intervals and variable rankings of HR values between the discovery and validation cohorts. The use of an integrated CytoScore partially compensated for individual marker variability by giving an overall assessment of the magnitude of the cytokine storm. Notable individual markers were associated with fatal outcome, namely

Angiopoietin-2, CXCL13 and RAGE. While Angiopoietin-2 was less strongly correlated with vRNA than RAGE, it appears of significant interest in severe COVID-19. This angiogenic factor has pro-inflammatory effects on the vascular endothelium, can disrupt vascular integrity and has been associated with ARDS (*44*). Angiopoietin-2 levels have been associated with severe COVID-19 (*45*), and might be a potential druggable target.

Antibody responses likely contribute to viral control in acute SARS-CoV-2 infection, and the negative associations we observed between plasma vRNA and SARS-CoV-2-specific antibody responses support this model. Although there were no global differences in antibody levels between the critical and non-critical groups, fatality was differentially association with humoral responses. Mortality was overrepresented among patients whom, at DSO11, had low RBD-specific IgG and low total Spike-binding Ig, although not in those with low RBD-specific IgM response. As only the IgG isoform among RBD-specific antibodies is lower in deceased patients, there may be a disruption in B cell functions requiring T-cell help, like class-switching to IgG, possibly linked to inadequate T follicular helper (T_FH_) and/or germinal centers (GC) disruption (*46*). CXCL13 is a key chemokine for recruitment to the GC of T_FH_ and B cells (*47*), and plasma CXCL13 is a marker of GC activity (*48*). As such, the positive associations of CXCL13 levels with vRNA loads and fatality risk, and the inverse correlation of CXCL13 levels with antibody responses, may seem paradoxical, but high amounts of circulating CXCL13 might disrupt the dynamics of B cell recruitment to the GC. In addition, heightened systemic inflammation can impaired development of adaptive immunity (*49, 50*). These mechanisms may explain the reduced RBD-specific IgG in patients who succumb to their infection.

Defective early ADCC responses were also significantly associated with fatality, whereas we found only a non-significant trend for neutralization capacity. These observations support that Fc-mediated functions could be important in controlling SARS-CoV-2, in line with recent reports showing that compromised Fc receptor binding strongly correlated with COVID-19 mortality(*27*), and Spike-specific humoral responses, including higher Fc-effector functions, were enriched among survivors (*51*). Furthermore, antibodies with intact Fc-effector functions were required for optimal protection against infection and correlated with decreased viral loads in animal models (*52, 53*).

The significant interactions we observed between a number of the features measured are compatible with different, non-mutually exclusive mechanisms. Poor development of protective antibody responses may allow persistently high levels of viral replication, which in turn will lead to a cytokine storm. Conversely, high cytokine levels, perhaps driven by systemic vRNA, may disrupt adaptive immune responses. Although our observational study does not allow addressing the question of causation between the immunovirological alterations observed, these measurements can be useful tools to understand heterogeneity in disease trajectories and response to therapy, particularly in the context of large, well-controlled randomized controlled trials. High viral loads and low levels of SARS-CoV-2-specific IgG may be mitigated through antivirals, monoclonal antibodies or convalescent plasma therapy with high IgG content. People with high levels of selected cytokines may benefit the most from targeted immunotherapies. Recent trials have already resulted in marked improvement in clinical patient care. As our study has been conducted on patients hospitalized during the first COVID-19 wave in spring and early summer 2020, it will be important to assess how t interventions that only recently became part of standard clinical care, as well as future therapeutic strategies, affect the potential of such immunovirological monitoring not only to predict outcome, but potentially to individualize patient management.

## Data Availability

All data is provided in the manuscript.
Further information and requests for resources and reagents should be directed to the Lead Contact, Kaufmann Daniel

## ACKNOWLEDGMENTS

For sample collection at the CHUM, we would like to thank Dounia Boumahni, Fatna Benettaib, Ali Ghamraoui, Boaz Lahav, Pascale Arlotto, Nakome Nguissant, Marc Messier-Peet, Stéphanie Matte and Martine Lebrasseur. For sample processing at the CRCHUM, we acknowledge Gloria Ortega-Delgado, Mélanie Laporte, Caroline Dufour, Isabelle Turcotte, Syllah Mohammed and Natalia Zamorano. We thank Dr. Stefan Pöhlmann (Georg-August University, Germany) for the plasmid coding for SARS-CoV-2 S glycoproteins and Dr. M. Gordon Joyce (U.S. MHRP) for the monoclonal antibody CR3022. For sample collection and processing at JGH, we thank Meriem Bouab, Danielle Henry, Zaman Afrasiabi, Hershlee Vernet, Branka Vulesevic, Nardin Rezk, Nofar Kimchi, Chris Tselios, Charlotte Guzman, Louis Petitjean, Xiaoqing Xue, Maureen Oliveira and Bluma Brenner. For chart review at JGH, we also thank Yara Moussa, Olumide Adeleye, Noor Mamlouk, Tala Abdullah, Michael Palayew and Biswarup Ghosh. For administration at JGH, we thank Vincenzo Forgetta, Darin Adra, Jonathan Afilalo and Marc Afilalo. Finally, we also thank the flow cytometry and NC3 platforms at the CRCHUM.

## AUTHORS CONTRIBUTION

<u>Conceptualization:</u> E.B.R, D.E.K, N.C, A.F <u>Data Curation</u>: E.B.R, N.B, C.O, C.L, D.E.K, R.M.R,

G.B.L, D.M, T.N, S.Z <u>Formal analysis</u>: E.B.R, S.P.R, P.G, G.M.B, A.D, G.B.B, A.P, R.G, J.R,

D.E.K. <u>Funding acquisition</u>: D.E.K, A.F, N.C, N.A, A.P, C.L, J.B.R, M.T, M.D.

<u>Investigation/experiments:</u> S.P.R, P.G, E.B.R, G.B.B, A.P, M.B, F.P, R.G, A.L, J.R, J.P, G.G, J.N,

M.N, G.S, H.M, C.B, J.D.D, M.B, G.G.L <u>Biostatistical methodology:</u> A.D, G.M.B <u>Patient</u>

<u>recruitment and cohort administration:</u> N.B, D.M, L.L, A.P, M.D, J.B.R, M.C, D.E.K <u>Visualization:</u>

E.B.R, G.M.B, A.D <u>Supervision:</u> M.C, M.T, N.C, A.F, D.E.K. <u>Writing – original draft:</u> E.B.R, D.E.K.

<u>Writing – review and editing</u>: all authors.

Every author has read, edited and approved the final manuscript.

## STAR METHODS

### Participants and samples

SARS-CoV-2 positive patients admitted to the Centre Hospitalier de l’Université de Montréal (CHUM or the Jewish General Hospital (JGH) were recruited into the *Biobanque Québécoise de la COVID-19* (BQC19). Samples from CHUM made up the discovery cohort, and samples from JGH were the validation cohort. Blood draws were performed at baseline and when possible, at Day 2 (± 3 days) and Day 7 (± 3days) after enrollment. The study was approved by the respective IRBs and written, informed consent obtained from all participants or, when incapacitated, their legal guardian before enrollment and sample collection. Blood draws were also performed on 50 asymptomatic, NSW PCR negative uninfected controls (UC).

COVID-19 hospitalized patients were stratified based on severity of respiratory support at the DSO11 timepoint: critical patients required mechanical ventilation (endotracheal, non-invasive ventilation, extracorporeal membrane oxygenation - ECMO), and non-critical patients, encompassing patients with moderate disease required no supplemental oxygen and patients with severe disease required nasal cannula for oxygen. Mortality was followed up to 60 days. Medical charts were reviewed by two physicians for data collection on demographics, co-morbidities, risk factors, severity state, time of infection, etc (see Table 1). Median age and range for UC cohort was 37 (32–46), and 30 individuals were males (60%).

### Cell lines

293T human embryonic kidney cells (obtained from ATCC) were maintained at 37°C under 5% CO_2_ in Dulbecco’s modified Eagle’s medium (DMEM) (Wisent) containing 5% fetal bovine serum (VWR) and 100 μg/mL of penicillin-streptomycin (Wisent). The 293T-ACE2 were previously reported(*25*).

For the generation of CEM.NKR CCR5+ cells stably expressing the SARS-CoV-2 Spike protein, transgenic lentiviruses were produced in 293T using a third-generation lentiviral vector system, as previously reported(*54*). Briefly, 293T cells were co-transfected with two packaging plasmids (pLP1 and pLP2), an envelope plasmid (pSVCMV-IN-VSV-G) and a lentiviral transfer plasmid coding for a GFP-tagged SARS-CoV-2 Spike (pLV-SARS-CoV-2 S C-GFPSpark tag) (Sino Biological). Supernatant containing lentiviral particles was used to transduce CEM.NKR CCR5+ cells in presence of 5μg/mL polybrene. The CEM.NKR CCR5+ cells stably expressing SARS-CoV-2 Spike (GFP+) were sorted by flow cytometry.

### Quantification of SARS-CoV2 RNA

Absolute copy numbers of SARS-CoV-2 RNA (N region) in plasma samples were measured by real time PCR. Total RNA was extracted from 230 μL of plasma collected on ACD using the QIAamp Viral RNA Mini Kit (Qiagen Cat. No. 52906), according to manufacturer’s instructions. Real-time PCR reactions were performed in 384-well plates using the TaqPath 1-Step Multiplex Master Mix (No ROX) (Applied Biosystems Cat. No. A28521) on a QuantStudio 5 instrument. Two master reaction mixes with specific primers and probes were prepared for quantification of N gene from SARS-CoV-2 and 18S (as a control for efficient extraction and amplification). The RT-qPCR reaction was performed in a volume of 15 μL including 3.75 μL of Taqpath Master Mix, 1.88 μL of a 8X mix of primers/probe (providing a final concentration of each primer of 400 nM and probe of 200 nM), 4.38 μL of H2O and 5 μL of the RNA extracts, controls or standards. The cycling parameters for qPCR were 53°C for 10 min, 95°C for 2 min and then 95°C for 3 s and 60°C for 30 s for 45 cycles. N SARS-CoV-2 quantifications were performed in quadruplicate and 18S measurements were performed in duplicates. The sequences of the primers and probes were as follows: N_IDT_F, 5’ CGTACTGCCACTAAAGCATACA 3’; N_IDT_R, 5’GCGGCCAATGTTTGTAATCAG3’; N_IDT_P, 5’AGACGTGGTCCAGAACAAACCCAA 3’ (HEX-ZEN-IABQ); 18S-F, 5’GTAACCCGTTGAACCCCATT3’; 18S-R, 5’CCATCCAATCGGTAGTAGCG3’; 18S-HEX, 5’CTTTGTACACACCGCCCG3’ (HEX-ZEN-IABQ). A positive and no-template controls were included in all experiments.

To obtain absolute copy numbers of N SARS-CoV-2 transcripts, in vitro transcribed RNA standards were generated. Linear templates for in vitro transcription containing a T7 promoter upstream of the complete N sequence were generated by PCR from a commercially available plasmid (2019-nCoV-N_positive plasmid – IDT Cat. No. 10006625). The PCR reaction was performed in a total volume of 40 μL: 0.5 μL of Taq Polymerase (Invitrogen Cat. No.18038042), 5 μL of 10X Reaction Mix, 3 μL of MgCl2 (50mM), 1.5 μL of dNTP (10mM), 2.5 μL of each primer (10 μM, providing a final concentration of each primer of 500 nM), 34 μL of H2O, and 1 μL of the plasmid at 1ng/μL. The sequences of the primers are as follows: M13-T7_Ngene_F: 5’GTAAAACGACGGCCAGTTAATACGACTCACTATAGGGATGTCTGATAATGGACCCCAAAAT3’; M13 Reverse, 5’CAGGAAACAGCTATGAC3’. The cycling parameters were 94°C for 3 min and then 94°C for 45 s, 55°C 30 s and 72°C for 90 s for 30 cycles of amplification, with a final extension at 72°C for 10 min. The PCR product was purified using the QIAquick PCR purification kit according to the manufacturer’s protocol (Qiagen Cat. No. 28104). RNA transcripts were produced in vitro using the MEGAscript T7 Transcription Kit (Invitrogen Cat. No. AM1333). The transcription reaction was performed in a total volume of 20 μL: 2 μL of 10X Reaction Buffer, 2 μL of each rNTP, 2 μL of Enzyme Mix, and 0.5-2 pmol M13-flanked DNA template. The reaction was performed at 37°C for 4 hrs, then 1 μL of TURBO DNase was added and incubated for an additional 15 min at 37°C. RNA transcripts were then purified using the RNeasy mini kit according to the manufacturer’s protocol (Qiagen Cat. No. 74104). Purified RNA N transcripts (1328 bp) were quantified by Nanodrop and the RNA copy numbers were calculated using the ENDMEMO online tool (http://www.endmemo.com/bio/dnacopynum.php). Aliquots of 1011 copies/μL were stored at -80°C. For each qPCR batch, one aliquot was thawed and six serial dilutions were prepared to generate a standard curve (500,000 to 5 copies per PCR well).

### Measurements of plasma analytes by beads array

Never-thawed plasma aliquots were thawed at RT and SARS-CoV-2 virus was inactivated using 1% Triton-X100 for 2 hrs at RT. After inactivation, measurements were performed in duplicates using a customized Human Magnetix Luminex Assay (LXSAHM-26, R&D, see supplemental material for full list of analytes), as per manufacturer’s instructions. Plates were acquired using a Bio-plex 200 array system (Bio-Rad Laboratories) for CHUM samples, or MagPix® System (Luminex) for samples from JGH.

For analysis of analytes, raw fluorescence intensity values were first manually background-subtracted. Concentrations were extrapolated using each plate’s individual standard curve for all analytes using R package nCal with bcrm models(*55*). RAGE values for plate 1 were imputed using k-nearest neighbour analysis using the preProcess function in R package caret (Max Kuhn (2020). caret: Classification and Regression Training. R package version 6.0-86. https://CRAN.R-project.org/package=caret). PCA plot was generated using R package factoextra and the fviz_pca_biplot() function. Heatmaps were generated using R package pheatmap (Raivo Kolde (2019) pheatmap: Pretty Heatmaps. R package version 1.0.12. https://CRAN.R-project.org/package=pheatmap) and correlation plots from R package corrplot (Taiyun Wei and Viliam Simko (2017). R package “corrplot”: Visualization of a Correlation Matrix) (Version 0.84, available from https://github.com/taiyun/corrplot). Statistical analyses were run using the multcomp and Hmisc R packages(*56*) (R package version 4.4-1. https://CRAN.R-project.org/package=Hmisc).

Some cytokines and tissue damage markers were at very low concentrations, and the quantification platform we used was not sensitive enough to reliably them in most samples. As such, analytes with extrapolated values >90% and negative values>15% were identified by ⊘ in Figures 2, 3 and S2.

### CytoScore

For k analytes (n=26), the CytoScore for each sample was calculated as follows

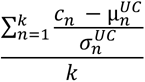

where

*cn* is the concentration for analyte n,

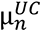 is the mean concentration of uninfected control samples for analyte n

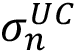 is the standard deviation of uninfected control samples for analyte n

### Plasma and antibodies

Previously thawed and refrozen (1 cycle) plasma was thawed and heat-inactivated for 1 hr at 56 °C and stored at -80°C until ready to use in subsequent experiments. Plasma from uninfected donors were used as negative controls and used to calculate the seropositivity threshold in our ELISA and flow cytometry assays. The monoclonal antibody CR3022 was used as a positive control in ELISA assays and was previously described (5–7). Horseradish peroxidase (HRP)-conjugated antibody specific for the Fc region of human IgG (Invitrogen), the Fc region of human IgM (Jackson ImmunoResearch Laboratories, inc.) or the Fc region of human IgA (Jackson ImmunoResearch Laboratories, inc) were used as secondary antibodies to detect antibody binding in ELISA experiments. Alexa Fluor-647-conjugated goat anti-human IgG (H+L) Abs (Invitrogen) were used as secondary antibodies to detect plasma binding in flow cytometry experiments.

### RBD-specific ELISA

The SARS-CoV-2 RBD assay used was recently described(*25, 34*). Briefly, recombinant SARS-CoV-2 S RBD proteins (2.5 μg/mL), or bovine serum albumin (BSA) (2.5 μg/mL) as a negative control, were prepared in PBS and were adsorbed to plates (MaxiSorp Nunc) overnight at 4°C. Coated wells were subsequently blocked with blocking buffer (Tris-buffered saline [TBS] containing 0.1% Tween20 and 2% BSA) for 1h at room temperature. Wells were then washed four times with washing buffer (Tris-buffered saline [TBS] containing 0.1% Tween20). CR3022 mAb (50ng/mL) or plasma from SARS-CoV-2-infected (1/250 dilution to detect IgA and IgM binding; 1/500 dilution to detect IgG binding) or uninfected donors (1/250 dilution) were prepared in a diluted solution of blocking buffer (0.1 % BSA) and incubated with the RBD-coated wells for 90 mins at room temperature. Plates were washed four times with washing buffer followed by incubation with secondary Abs prepared in a diluted solution of blocking buffer (0.4% BSA) for 1h at room temperature, followed by four washes. HRP enzyme activity was determined after the addition of a 1:1 mix of Western Lightning oxidizing and luminol reagents (Perkin Elmer Life Sciences). Light emission was measured with a LB942 TriStar luminometer (Berthold Technologies). Signal obtained with BSA was subtracted for each plasma and was then normalized to the signal obtained with CR3022 mAb present in each plate. The seropositivity threshold was established using the following formula: mean of all COVID-19 negative plasma + (3 standard deviation of the mean of all COVID-19 negative plasma).

### Flow cytometry analysis of cell-surface staining

Using the standard calcium phosphate method, 10 μg of SARS-CoV-2 Spike expressor and 2 μg of a green fluorescent protein (GFP) expressor (pIRES-GFP) were transfected into 2 × 10^6^ 293T cells. At 48 hrs post transfection, 293T cells were stained with plasma from SARS-CoV-2-infected or uninfected individuals (1/250 dilution). The percentage of transfected cells (GFP+ cells) was determined by gating the live cell population based on the basis of viability dye staining (Aqua Vivid, Invitrogen). Samples were acquired on a LSRII cytometer (BD Biosciences, Mississauga, ON, Canada) and data analysis was performed using FlowJo vX.0.7 (Tree Star, Ashland, OR, USA). The seropositivity threshold was established using the following formula: mean of all COVID-19 negative plasma + (3 standard deviation of the mean of all COVID-19 negative plasma + inter-assay coefficient of variability).

### Virus neutralization assay

293T-ACE2 target cells were infected with single-round luciferase-expressing pseudoparticles bearing the SARS-CoV-2 Spike in presence of patient plasma. Briefly, 293T cells were transfected by calcium phosphate method with the lentiviral vector pNL4.3 R-E-Luc (NIH AIDS Reagent Program) and a plasmid encoding for SARS-CoV-2 Spike at a ratio of 5:4. Two days post-transfection, cell supernatants were harvested and stored at -80°C until use. 293T-ACE2 target cells were seeded at a density of 1×10^4^ cells/well in 96-wells luminometer-compatible tissue culture plates (Perkin Elmer) 24 hrs before infection. Recombinant viruses in a final volume of 100μl were incubated with the indicated plasma dilutions (1/50; 1/250; 1/1250; 1/6250; 1/31250) for 1 hr at 37°C and were then added to the target cells followed by incubation for 48 hrs at 37°C; cells were lysed by the addition of 30μl of passive lysis buffer (Promega) followed by one freeze-thaw cycle. An LB942 TriStar luminometer (Berthold Technologies) was used to measure the luciferase activity of each well after the addition of 100 μL of luciferin buffer (15mM MgSO4, 15mM KPO4 [pH 7.8], 1mM ATP, and 1mM dithiothreitol) and 50 μL of 1mM d-luciferin potassium salt (Thermo Fisher Scientific). The neutralization half-maximal inhibitory dilution (ID50) represents the plasma dilution to inhibit 50% of the infection of 293T-ACE2 cells by pseudoviruses bearing the SARS-CoV-2 S protein.

### ADCC assay with SARS-CoV-2 Spike expressing cells

For evaluation of antibody-dependent cellular cytotoxicity (ADCC), parental CEM.NKr CCR5+ cells were mixed at a 1:1 ratio with CEM.NKr. Spike cells. These cells were stained for viability (AquaVivid; Thermo Fisher Scientific, Waltham, MA, USA) and cellular (cell proliferation dye eFluor670; Thermo Fisher Scientific) dyes to be used as target cells. Overnight rested PBMCs were stained with another cellular marker (cell proliferation dye eFluor450; Thermo Fisher Scientific) and used as effector cells. Stained target and effector cells were mixed at a ratio of 1:10 in 96-well V-bottom plates. Plasma from COVID-19 or uninfected individuals (1/500 dilution) or monoclonal antibody CR3022 (1 µg/mL) were added to the appropriate wells. The plates were subsequently centrifuged for 1 min at 300xg, and incubated at 37°C, 5% CO_2_ for 5 to 6 hrs before being fixed in a 2% PBS-formaldehyde solution. ADCC was calculated by gating on Spike-expressing live target cells and using the formula:

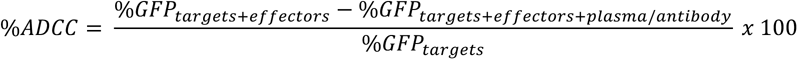

%ADCC obtained with plasma was further normalized to the %ADCC measured with CR3022. All samples were acquired on an LSRII cytometer (BD Biosciences) and data analysis was performed using FlowJo v10.0.7 (Tree Star). The specificity threshold was established using the following formula: mean of all COVID-19 negative plasma + 3 standard deviation of the mean of all COVID-19 negative plasma.

### Statistical analyses and multivariate models

#### Methods for univariate models

The association between of measured variables and time to death was analyzed by Cox Proportional Survival Hazard. The dependent variable in all survival analyses was time to death during the follow-up, measured in days. Subjects were censored upon reaching 60 days of follow-up (no patients withdrew within this timeframe). The time 0 was defined as a day of symptoms onset. Univariate Cox Proportional Hazard regression was used to determine the association plasma analytes and all-cause mortality at DSO60 for all COVID-19+ patients, as well as critical patients’ subgroup only. Analytes were log-transformed when they naturally followed exponential distribution, for example vRNA and cytokines. Next, the estimated survival proportions at any given point in time for a undetectable (when applicable), low lower interquartile range level of detectable) or high (upper interquartile range of detectable) were extracted from Cox models(*29*) and presented in the graphical form(*29*).

#### Part 1. Multivariate Cox model

Potential risk factors were grouped in 3 categories: i) vRNA, ii) 26 cytokine variables and iii) 6 antibody variables. Model building was performed in three steps. In the first step, univariate models for risk factor of death by DSO60 were developed, one for each of the covariates in the category; only risk factors p value <0.05 were retained. For the second category of 26 cytokines, an additional criterion of variable selection was applied to ensure the quality of the measurements: the cytokines with extrapolated values >90% and negative values>15% were excluded for future investigation. These exclusion criteria were added as the quantification platform we used was not sensitive enough to reliably quantify some low-concentration analytes, and we wanted to rely on analytes which are well quantified for our multivariate model. 19 cytokines out of 26 were satisfied these criteria. In the second step, categories for which more than one variable had been retained in the first step were focused on; then the stepwise Cox model selection based on the Bayesian Information Criterion (BIC) was used to obtain the most-parsimonious model (lowest value) for each of these categories. This penalized likelihood criterion selects the best variable at predicting data, then adds one additional variable at a time while accounting for potential overfitting, in the end only selecting the multivariate model with the lowest BIC value, i.e. the most parsimonious. In addition, to keep the risk of overfitting low, no more than six predictor parameters were entered in the multivariate model for our sample of 61 patients(*57, 58*).

In the third step, all variables retained in the second step were considered; then the BIC was used to obtain a global parsimonious model. Based on the literature (*59*), age and sex are associated with the mortality for COVID-19 patients, however in the small homogeneous sample in might be hard to detect these relations. Thus, in each model Age and Sex covariates were forced in the multivariate model regardless their significance. Potential interactions between each covariate with age and sex were tested to verify if the effect is consistent across different age and between sex. Potential presence of multicollinearity was assessed by calculating the variance inflation factor (VIF) for each variable. This allowed us to identify and treat in separate models subsets of covariates which were highly correlated.

#### Part 2. Time dependent ROC curve

To evaluate predictive accuracy of survival models the time-dependent receiver operator characteristic (ROC) curves for right-censored data (*60*) were calculated, compared across different Cox models and presented in the graphical form. The inverse probability of censoring weighting technique (IPCW) was used for estimating time-dependent ROC curves (*61*). The area under the curve (AUC) was examined at 60 days as well at all distinct event times by plotting the AUC curve and the 95% confidence limits over time. The day 48 corresponds the last event (fatality) day in the discovery cohort.

#### Part 3. Independent cohort validation

All final multivariate Cox models were reassessed in the validation cohort by executing independently the multivariate models with the same list of variables obtained, in the discovery cohort, in steps 2 and 3. Then, using the same approach described above, the time dependent ROC curves were evaluated in validation dataset to validate our finding.

