## Supplemental Materials for "Integrated immunovirological profiling validates plasma SARS-CoV-2 RNA as an early predictor of COVID-19 mortality"

Figure S1. Study design.

Figure S2. Inflammatory cytokines, chemokines and markers of tissue damage are increased in critical cases of COVID-19.

Figure S3. Association of poor outcome with low RBD-specific IgG titers is maintained in the critical COVID-19 group.

Figure S4. Reproducibility of predictive accuracy for mortality in the validation cohort.

Figure S5. Predictive accuracy of immunovirological markers, in the discovery and validation cohorts.

Table S1: Full list of analytes measured in plasma by beads arrays

**Figure S1. Study design. A)** Study Design. **B**) Survival curve in entire discovery cohort based on days since symptom onset. **C**) Kaplan-Meier analysis of survival in Non-critical (blue) compared to Critical (red) subgroups, whose disease severity was assessed at DSO11. Curves compared using Log-rank (Mantel-Cox) test. n = 61 COVID-19 patients (13 fatalities).

**Figure S2. Inflammatory cytokines, chemokines and markers of tissue damage are increased in critical cases of COVID-19. A-D)** Comparison of cytokine concentrations between critical COVID-19, non-critical COVID-10 and UC. Cytokines and markers of tissue damage grouped according to differential detection: **A)** Greatest in critical (Crt), but also higher in non-critical (N-Crt) compared to UC; **B)** Similar between UC and non-critical, but greater in critical COVID-19; **C)** No differences between all three groups; **D)** Greater in COVID-19 compared to UC, but similar between non-critical and critical. **E)** Correlation matrix of all 26 plasma analytes and CytoScore (see methods for details on CytoScore). . Color of circle represents Spearman R value (red = 1, blue = -1) and respective p values are represented by * within circles (p < 0.05 = *; p < 0.01 =** ; p<0.001=*** ; p<0.0001=****). **FG)** Correlation of plasma vRNA and plasma concentration of **F)** IL-6 or **G)** RAGE (pg/mL). **H)** Comparison of CytoScore between aviremics (<13 vRNA copies/mL) and viremics (≥ 13 copies/mL). Mann-Whitney test. A-D) Kruskall-Wallis with Dunn's multiple comparisons test. For A-D, F-H, color-coded dots represent severity of the patient at DSO11 (red = critical, blue = non-critical, green=UC). For A-E, cytokines with titles annotated by ∅ are poorly detected (see methods for details). n = 61 COVID-19 subjects (13 mortalities) and 43 UC.

**Figure S3. Association of poor outcome with low RBD-specific IgG titers is maintained in the critical COVID-19 group. A-C)** Modelisation of the hazard ratio of patients with high (orange) or low (purple) **A)** RBD-specific IgG, **B)** Spike-specific Ig or **C)** ADCC activity in critical COVID-19 patients. n = 29 Critical COVID-19 cases (11 mortalities).

**Figure S4. Reproducibility of predictive accuracy for mortality in the validation cohort**. **A-C)** Time-dependent ROC curves measured within the validation cohort for **A)** plasma vRNA, age and sex; **B)** Cytokines and tissue insult markers or **C)** anti-SARS-CoV-2 antibody responses. **D**) Table summarizing time-dependent AUC for representative variables per category in discovery cohort. Values are AUC (95%CI), and AUC at p<0.05 are in bold. AUC values given either not adjusted (only variable(s) listed) or adjusted (age and sex). AUC values are given at 30 days or 60 days after symptom onset. Maximum AUC column gives the best prediction accuracy of the variable (measured at DSO11), and DSO for maximum is the date since symptom onset at which that maximum AUC was achieved. A-C) Legends with color-coded variables are on the bottom left of panels, and values in italic are the AUC values associated to the variable or model. n = 83.

**Figure S5. Predictive accuracy of immunovirological markers, in the discovery and validation cohorts**. **A d**) Concept of time-dependent AUC changes. To observe changing accuracy overtime, AUCs for a given measurement were plotted against time to death. AUC values closer to 1 (top) have better predictive accuracy; AUC values close to 0.5 have poor prediction accuracy. In this example, measurement A maintained the greatest prediction accuracy throughout time, whereas C > B before DSO30, then B > C after DSO30. **BC)** Time-dependent AUC of plasma vRNA, age and sex for **B**) discovery cohort or **C**) validation cohort; **DE)** Time-dependent AUC of plasma cytokines and tissue damage markers for **D**) discovery cohort or **E**) validation cohort; **FG)** Time-dependent AUC of SARS-CoV-2 antibody responses for **F**) discovery cohort or **G**) validation cohort. **HI**) Time-dependent AUC for top measurements captured by multivariate model analysis in the **H**) discovery cohort or **I**) validation cohort. n = 61 for discovery cohort; 83 for validation cohort. Legends with color-coded variables are on the bottom left of panels, and values in italic are the AUC values associated to the variable. n = 61 for discovery cohort; 83 for validation cohort.

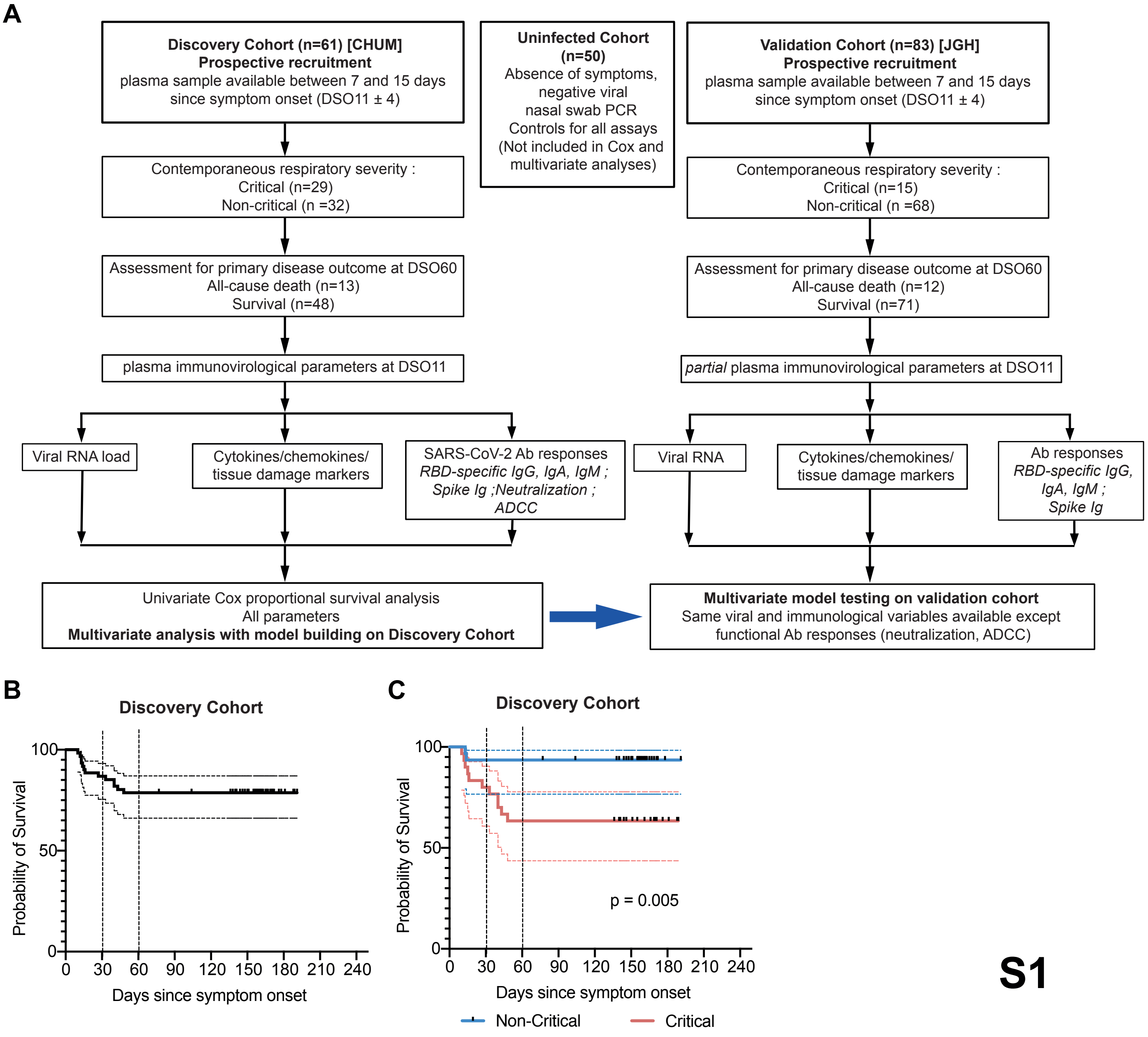
A
Discovery Cohort (n=61) [CHUM]
Prospective recruitment
plasma sample available between 7 and 15 days since symptom onset (DSO11 ± 4)
Uninfected Cohort (n=50)
Absence of symptoms, negative viral nasal swab PCR
Controls for all assays
(Not included in Cox and multivariate analyses)
Validation Cohort (n=83) [JGH]
Prospective recruitment
plasma sample available between 7 and 15 days since symptom onset (DSO11 ± 4)
Contemporaneous respiratory severity :
Critical (n=29)
Non-critical (n =32)
Contemporaneous respiratory severity :
Critical (n=15)
Non-critical (n =68)
Assessment for primary disease outcome at DSO60
All-cause death (n=13)
Survival (n=48)
Assessment for primary disease outcome at DSO60
All-cause death (n=12)
Survival (n=71)
plasma immunovirological parameters at DSO11
partial plasma immunovirological parameters at DSO11
Viral RNA load
Cytokines/chemokines/ tissue damage markers
SARS-CoV-2 Ab responses
RBD-specific IgG, IgA, IgM ; Spike Ig ;Neutralization ; ADCC
Viral RNA
Cytokines/chemokines/ tissue damage markers
Ab responses
RBD-specific IgG, IgA, IgM ; Spike Ig
Univariate Cox proportional survival analysis
All parameters
Multivariate analysis with model building on Discovery Cohort
Multivariate model testing on validation cohort
Same viral and immunological variables available except functional Ab responses (neutralization, ADCC)
B
Discovery Cohort
Probability of Survival
100
50
0
0 30 60 90 120 150 180 210 240
Days since symptom onset
C
Discovery Cohort
Probability of Survival
100
50
0
p = 0.005
0 30 60 90 120 150 180 210 240
Days since symptom onset
Non-Critical
Critical

S1

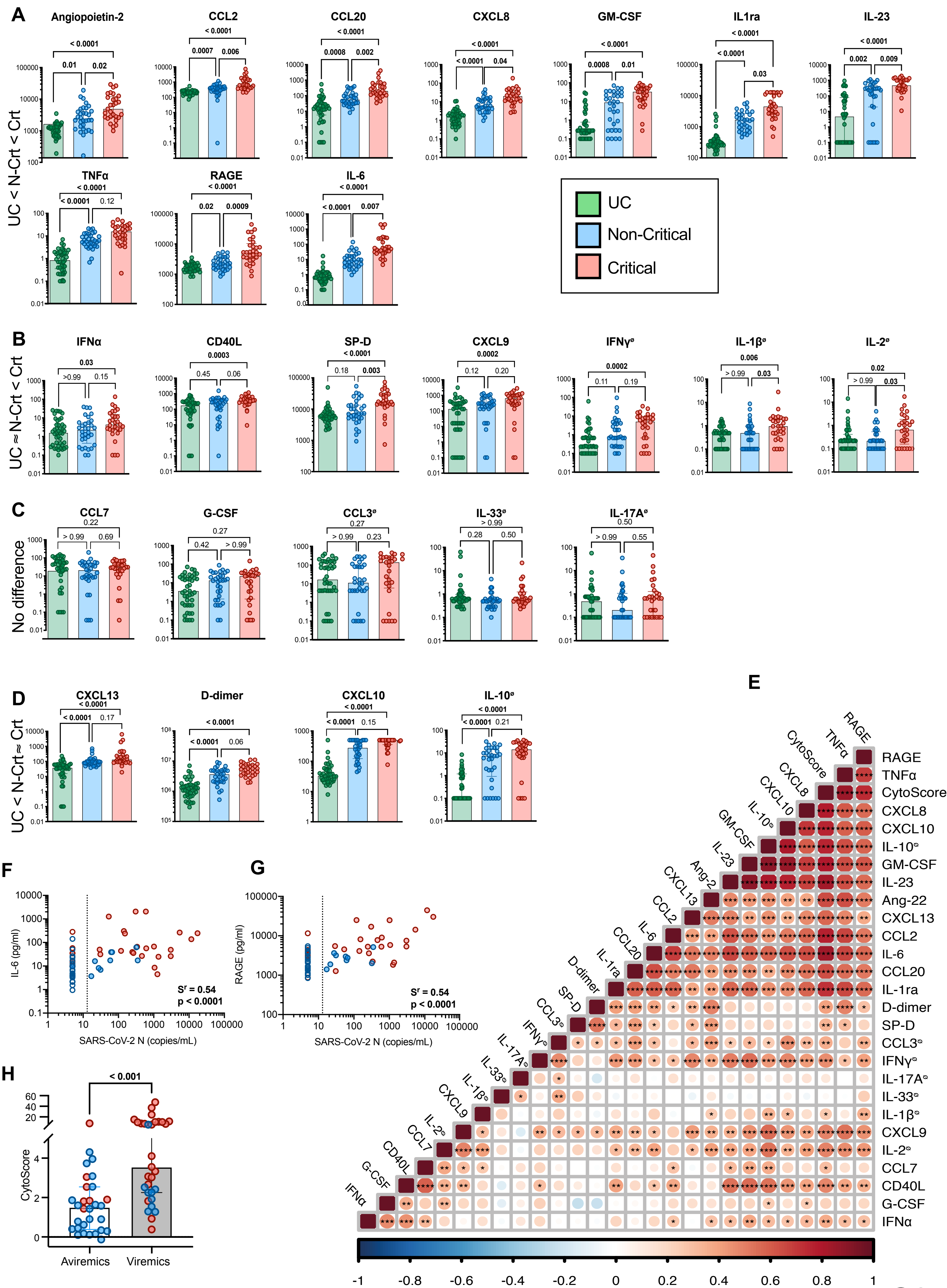
A
Angiopoietin-2
CCL2
CCL20
CXCL8
GM-CSF
IL1ra
IL-23
TNFα
RAGE
IL-6
UC < N-Crt < Crt
UC
Non-Critical
Critical
B
IFNα
CD40L
SP-D
CXCL9
IFNγ
IL-1β
IL-2
UC ≈ N-Crt < Crt
C
CCL7
G-CSF
CCL3
IL-33
IL-17A
No difference
D
CXCL13
D-dimer
CXCL10
IL-10
UC < N-Crt ≈ Crt
E
RAGE
TNFα
CytoScore
CXCL8
CXCL10
IL-10
GM-CSF
IL-23
Ang-2
Ang-22
CXCL13
CCL2
IL-6
CCL20
IL-1ra
D-dimer
SP-D
CCL3
IFNγ
IL-17A
IL-33
IL-1β
CXCL9
IL-2
CCL7
CD40L
G-CSF
IFNα
F
IL-6 (pg/ml)
SARS-CoV-2 N (copies/mL)
Sr = 0.54
p < 0.0001
G
RAGE (pg/ml)
SARS-CoV-2 N (copies/mL)
Sr = 0.54
p < 0.0001
H
CytoScore
< 0.001
Avirемics
Viremics

S2

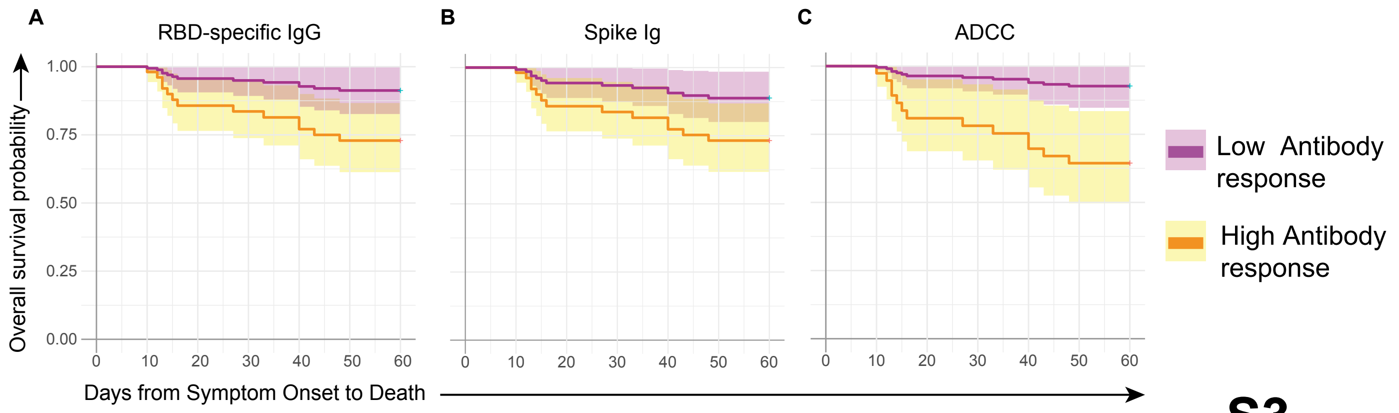
A
RBD-specific IgG
B
Spike Ig
C
ADCC
Overall survival probability
1.00
0.75
0.50
0.25
0.00
0
10
20
30
40
50
60
Days from Symptom Onset to Death
Low Antibody response
High Antibody response

**S3**

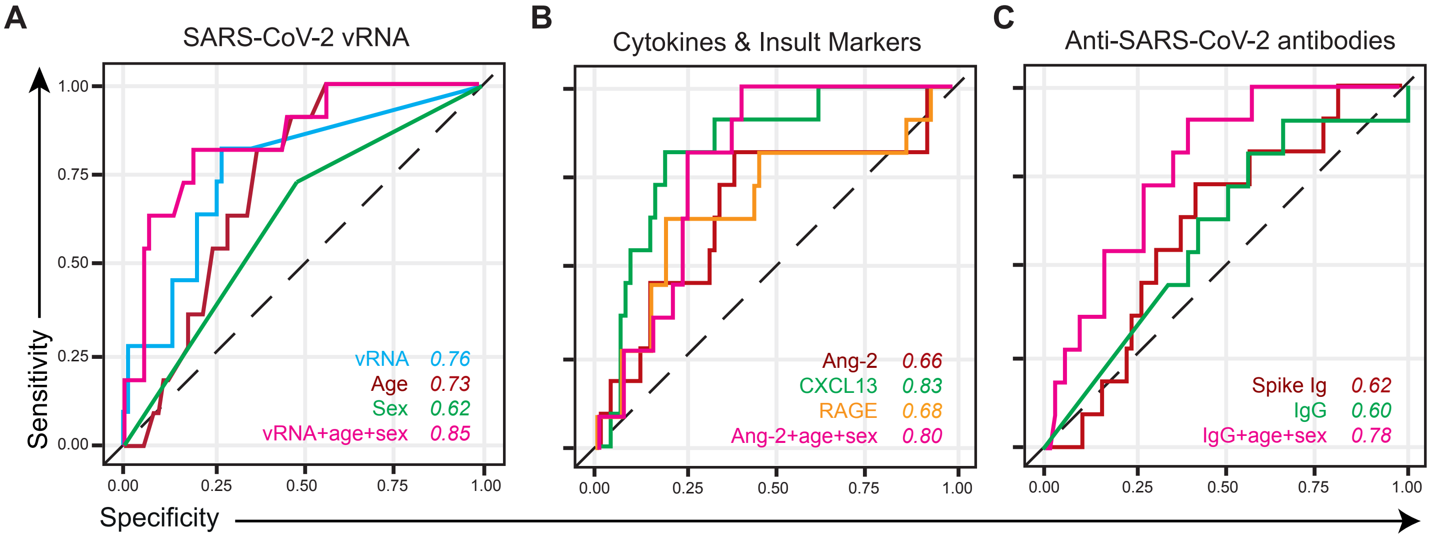


D

| AUC(95%CI) - Validation Cohort | | | | | | |
|---|---|---|---|---|---|---|
| Model | | 30 Days since Symptom Onset | 60 Days since Symptom Onset | Integrated AUC | Maximum AuC | DSO for Maximum AUC |
| vRNA | Not-adjusted | **0.73 (0.58, 0.88)** | **0.76 (0.60, 0.92)** | 0.82 | **0.91 (0.81, 1.00)** | 12 |
| | Adjusted (age+sex) | **0.88 (0.70, 1.00)** | **0.85 (0.67, 1.00)** | 0.88 | **0.92 (0.73, 1.00)** | 22 |
| Ang-2 | Not-adjusted | 0.63 (0.19, 1.00) | 0.66 (0.19, 1.00) | 0.64 | 0.95 (0.27, 1.00) | 8 |
| | Adjusted (age+sex) | 0.82 (0.33, 1.00) | 0.80 (0.32, 1.00) | 0.79 | 0.83 (0.30, 1.00) | 8 |
| IgG | Not-adjusted | 0.54 (0.17, 0.92) | 0.60 (0.18, 1.00) | 0.55 | **0.85 (0.58, 1.00)** | 22 |
| | Adjusted (age+sex) | **0.80 (0.54, 1.00)** | **0.78 (0.54, 1.00)** | 0.75 | 0.62 (0.22, 1.00) | 22 |
| vRNA + Ang-2 | Not-adjusted | 0.75 (0.26, 1.00) | 0.77 (0.26, 1.00) | 0.81 | 0.96 (0.30, 1.00) | 12 |
| | Adjusted (age+sex) | 0.86 (0.33, 1.00) | 0.85 (0.32, 1.00) | 0.89 | 0.94 (0.33, 1.00) | 12 |

**S4**

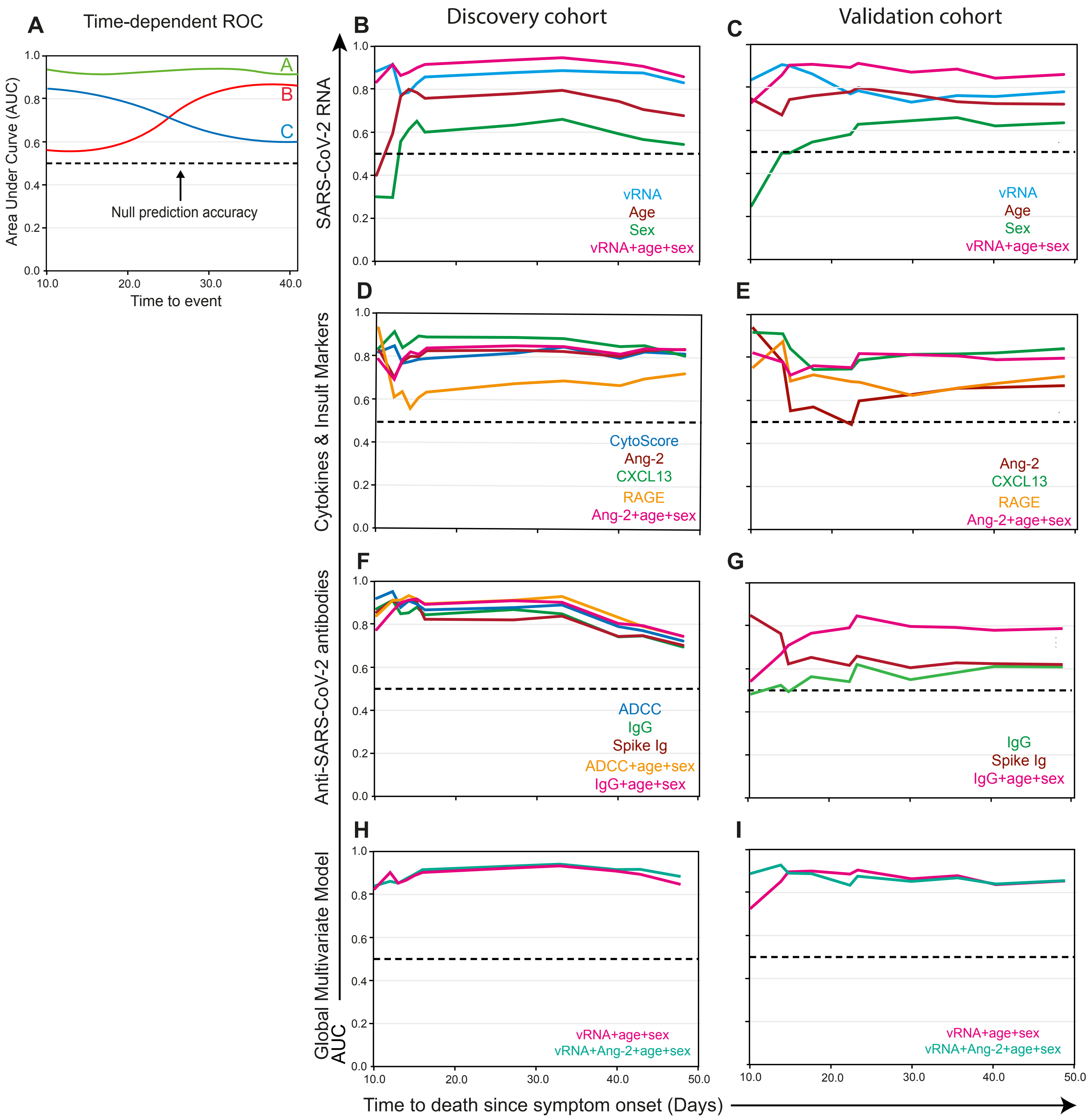

A
Time-dependent ROC
Area Under Curve (AUC)
Null prediction accuracy
Time to event
Discovery cohort
Validation cohort
B
C
SARS-CoV-2 RNA
vRNA
Age
Sex
vRNA+age+sex
D
E
Cytokines & Insult Markers
CytoScore
Ang-2
CXCL13
RAGE
Ang-2+age+sex
F
G
Anti-SARS-CoV-2 antibodies
ADCC
IgG
Spike Ig
ADCC+age+sex
IgG+age+sex
H
I
Global Multivariate Model
AUC
vRNA+age+sex
vRNA+Ang-2+age+sex
Time to death since symptom onset (Days)


**S5**

**Supplemental Table 1. Full list of analytes measured in plasma by beads arrays**

| Analyte | Bead Region |
|---|---|
| Angiopoietin-2 | 26 |
| CCL3/MIP-1 alpha | 35 |
| CCL20/MIP-3 alpha | 33 |
| CXCL9/MIG | 52 |
| CXCL13/BCA-1 | 28 |
| G-CSF | 54 |
| IFN$\alpha$ | 63 |
| IL-1$\beta$/IL-1F2 | 57 |
| IL-2 | 27 |
| IL-8/CXCL8 | 18 |
| IL-17/IL-17A | 43 |
| IL-33 | 14 |
| SP-D | 62 |
| CCL2/JE/MCP-1 | 25 |
| CCL7/MCP-3/MARC | 37 |
| CD40 Ligand/TNFSF5 | 74 |
| CXCL10/IP-10/CGR-2 | 21 |
| D-dimer | 43 |
| GM-CSF | 46 |
| IFN-$\gamma$ | 29 |
| IL-1ra/IL-1F3 | 30 |
| IL-6 | 13 |
| IL-10 | 22 |
| IL-23 | 76 |
| RAGE/AGER | 45 |
| TNF$\alpha$ | 12 |

Human Magnetic Luminex ® Assays, from R&D Systems (Biotechne)

Premixed Multiplex
Kit Catalog Numbers : LXSAHM-26

Kit Lot Number : L134818,
